# Influence of autozygosity on common disease risk across the phenotypic spectrum

**DOI:** 10.1101/2023.02.01.23285346

**Authors:** Daniel S. Malawsky, Eva van Walree, Benjamin M Jacobs, Teng Hiang Heng, Qin Qin Huang, Ataf H. Sabir, Saadia Rahman, Saghira Malik Sharif, Ahsan Khan, Maša Umićević Mirkov, 23andMe Research Team, Genes & Health Research Team, Danielle Posthuma, William G. Newman, Christopher J. Griffiths, Rohini Mathur, David A. van Heel, Sarah Finer, Jared O’Connell, Hilary C. Martin

## Abstract

Autozygosity is associated with rare Mendelian disorders and clinically-relevant quantitative traits. We investigated associations between F_ROH_ (fraction of the genome in runs of homozygosity) and common diseases in Genes & Health (N=23,978 British South Asians), UK Biobank (N=397,184), and 23andMe, Inc. We show that restricting analysis to offspring of first cousins is an effective way of removing confounding due to social/environmental correlates of F_ROH_. Within this group in G&H+UK Biobank, we found experiment-wide significant associations between F_ROH_ and twelve common diseases. We replicated the associations with type 2 diabetes (T2D) and post-traumatic stress disorder via between-sibling analysis in 23andMe (median N=480,282). We estimated that autozygosity due to consanguinity accounts for 5-18% of T2D cases amongst British Pakistanis. Our work highlights the possibility of widespread non-additive effects on common diseases and has important implications for global populations with high rates of consanguinity.

## Introduction

The prevalence of consanguinity, unions between related individuals, differs around the world, being relatively low in modern European-ancestry populations and higher in South Asia and the Middle East^1,2^. It often co-occurs with endogamy, unions between individuals from the same clan or social group^3–5^. These practices increase the rates of autozygosity i.e. stretches of homozygosity in the genome that are identical by descent. Autozygosity is known to increase the risk of rare congenital anomalies and recessive Mendelian disorders^6,7^, and has been associated with various other phenotypic outcomes, such as decreased height, fertility, and self-reported overall health^8,9^, and increased risk for complex diseases such Alzheimer’s disease^10^ and coronary artery disease (CAD)^11^. Notably, the prevalence of CAD and other complex diseases such as type 2 diabetes (T2D) is significantly higher in British South Asian individuals compared to White British individuals^12^. While this is undoubtedly partly due to social and environmental factors^12,13^ as well as differential additive genetic susceptibility at certain loci towards T2D in South Asians compared to white Europeans^14^, it is unclear whether higher rates of autozygosity could also contribute.

One mechanistic explanation for the association between autozygosity and certain traits and diseases is that autozygosity increases the chance of harbouring rare homozygous genotypes at damaging recessive variants, which are less effectively removed from the population by negative selection than dominantly-acting variants^15^. However, other potential explanations exist, such as the heterozygote advantage hypothesis, whereby heterozygosity for certain common variants leads to fitness advantages^15^, or, that the increased variance in additive genetic liability towards binary traits induces associations with autozygosity in the absence of non-additive effects^16^.

A challenging problem in assessing the relationship between autozygosity and phenotypes is that associations may be confounded by both population structure and the social circumstances in which consanguinity and endogamy are practised. For example, attempted replication of a previously-detected association with schizophrenia^17^ failed in reasonably powered cohorts^18,19^, suggesting potential confounding. In another example, it has been shown that a negative association in the Dutch population between depression and the fraction of the genome in runs of homozygosity (F_ROH_, a measure of autozygosity) was confounded by religious assortative mating, whereby religious individuals had higher F_ROH_ due to stricter endogamy^20^. Thus, the environmental and social factors that correlate with having related parents may produce spurious associations between autozygosity and disease phenotypes. However, experimental studies in nonhuman organisms that are free of social and environmental confounding show effects of autozygosity on several phenotypes^15,21–25^, suggesting that the observations in humans may be at least partially of genetic origin.

Here, we describe the patterns of consanguinity and examine the effect of autozygosity on disease risk across the phenotypic spectrum in two cohorts: the Genes & Health cohort, a population-based study of self-reported British Bangladeshi and British Pakistani individuals, and in UK Biobank individuals genetically inferred to have European and South Asian ancestries. We show that subsetting association analyses to highly consanguineous individuals better controls for social and environmental confounding. With this approach, we find significant associations between autozygosity and various diseases, several of which we replicate, using a different method, in a between-sibling analysis conducted in the 23andMe cohort. Via simulations, we show that these observed associations most likely stem from non-additive genetic effects. Our study quantifies the effect of autozygosity across the disease phenotypic spectrum for the first time, using a novel approach that addresses confounding, and highlights the possibility of widespread non-additive effects across diseases.

Since consanguinity is a sensitive topic for many communities, we have prepared a “Frequently asked questions” document for a lay audience in collaboration with the Community Advisory Board from Genes & Health, explaining the motivation for and results of our study, and placing them in wider context.

## Results

Our main analysis focuses on two cohorts, Genes & Health (G&H) and UK Biobank (UKB), both with electronic health record (EHR) data from primary and secondary care provided by the National Health Service (NHS) in England. G&H (n=44,190 with genetic and EHR data at the time of analysis) is a community based cohort of British Bangladeshi (65%) and Pakistani (35%) individuals recruited in London, Manchester and Bradford, UK. The dataset is reasonably representative of the background population, albeit likely with some over-sampling of individuals with chronic diseases since much of the recruitment was conducted in a primary care setting^26^. We additionally analysed individuals with genetically-inferred European and South Asian ancestries from UKB (UKBEUR and UKBSAS, respectively). We removed individuals for whom EHR data linkage was unavailable and one of each pair of individuals inferred to be third-degree relatives or closer, leaving 23,978 G&H individuals, 387,531 UKBEUR individuals, and 9,653 UKBSAS individuals. See Table 1 for descriptive statistics of the cohorts.

**Table 1.** Descriptive statistics of unrelated individuals in the G&H and UKB cohorts. F_ROH_ is the fraction of the genome in runs of homozygosity. The bottom row gives the number of individuals inferred to be offspring of first cousin/avuncular unions included in the “highly consanguineous” analyses described below. SD: standard deviation.

|  | G&H<br>(n=23,978) | UKBEUR<br>(n=387,531) | UKBSAS<br>(n=9,653) |
| --- | --- | --- | --- |
| % male | 47% | 46% | 54% |
| Age (years) -<br>mean (SD) | 44.9 (13.1) | 56.7 (8.0) | 53.4 (8.5) |
| Self-reported<br>ancestry | 65% Bangladesh<br>35% Pakistan | 94% Great Britain,<br>6% other European | 60% India, 21% Pakistan, 4%<br>Bangladesh, 15% other South Asian |
| $F_{ROH}$ mean (SD) | 0.0178 (0.025) | 0.0037 (0.0050) | 0.013 (0.022) |
| # “highly<br>consanguineous” | 4,034 (16.8%) | 977 (0.25%) | 754 (7.8%) |

### Consanguinity patterns in Genes & Health and UK Biobank

Given that G&H has high self-reported rates of consanguinity^26^ (9% in British Bangladeshi individuals, 36% in British Pakistani individuals), we first sought to genetically characterise consanguinity patterns in the cohort and compare them to UK Biobank. We applied a method we previously developed to infer an individual’s parental relatedness (PR) based on the distribution of runs of homozygosity (ROHs) in their genome^2^. The method infers ten classes of PR, some involving multiple generations of consanguinity (Methods). Rates of consanguinity (offspring of second cousins or closer) were very low in UKBEUR (2%), and higher in UKBSAS and G&H (29% and 33% respectively) (Figure 1a). In concordance with previous findings in G&H based on F_ROH_ distribution^26^, self-reporting of PR was imperfect (Figure 1b,c).

**Figure 1.**
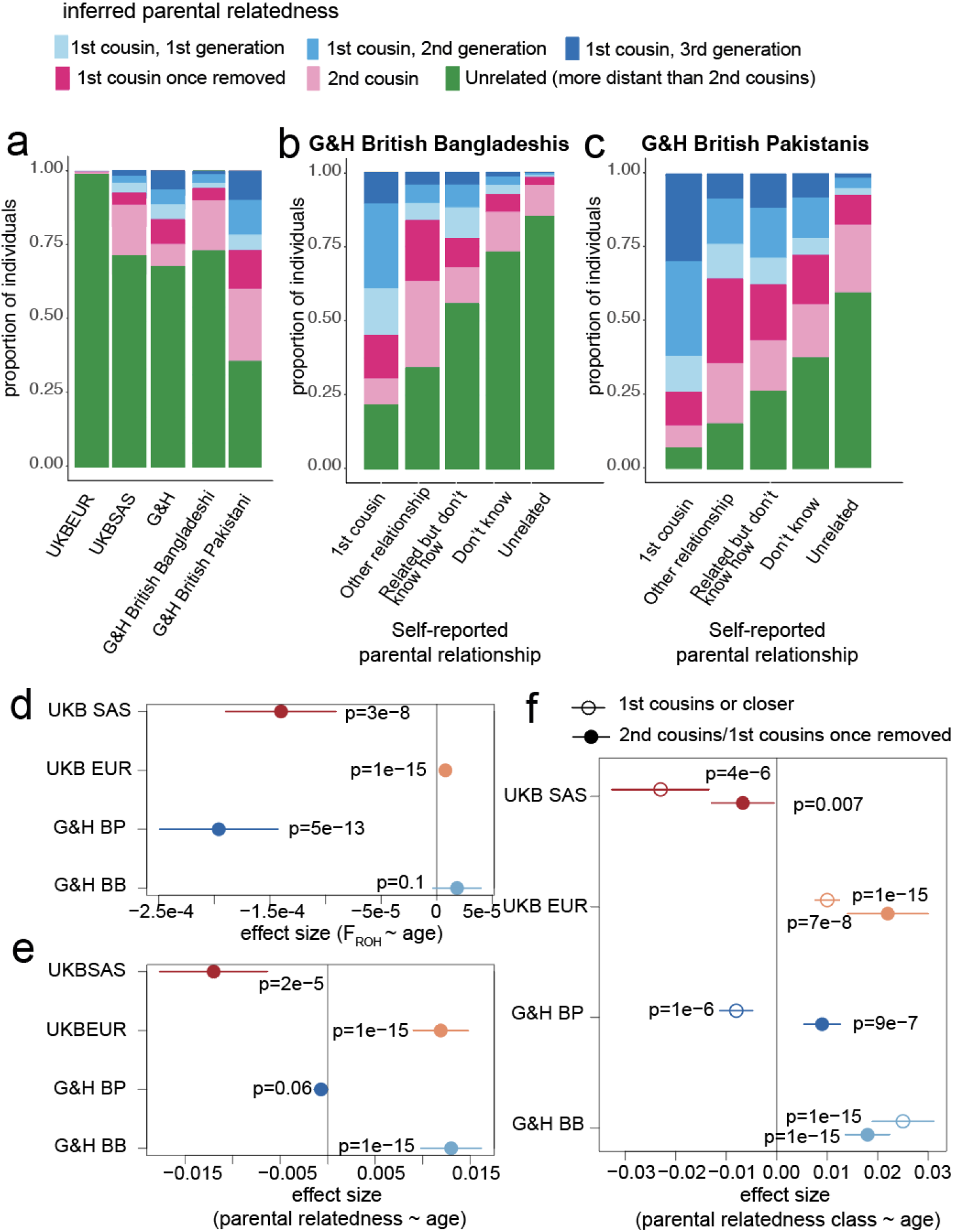
Patterns of parental relatedness (PR) in G&H and UKB. (a) Stacked bar plots showing genetically-inferred PR for the indicated groups. (b) and (c) Stacked bar plot showing genetically-inferred PR for G&H British Bangladeshi individuals and British Pakistani individuals respectively, stratified by self-reported PR. The inferred classes of PR include up to three generations of first cousin marriages, first cousin once removed, second cousin or unrelated. (d) Effect sizes of age on F_ROH_, inferred from linear regression, in the indicated groups. (e) Effect sizes of age on being genetically-inferred offspring of second cousins or closer, from logistic regression. (f) Effect sizes of age having the indicated class of PR, inferred from multinomial logistic regression. Lines indicated 95% confidence intervals. BB: British Bangladeshi; BP: British Pakistani.

Next, we explored whether the rate of consanguinity has been changing over time (Figure 1d,e,f). We replicated a recent finding ^27^ that, in UKBEUR, F_ROH_ significantly increases with age (Figure 1d). In contrast, F_ROH_ significantly decreases with age in G&H British Pakistani individuals but showed no significant association in G&H British Bangladeshi individuals (Figure 1d). In UKBEUR and G&H British Bangladeshi individuals, age was significantly positively associated with rates of both first cousin or closer PR and of first cousins once removed/second cousin PR (Figure 1f). In G&H British Pakistani individuals, although there is no significant overall change in the rate of PR (i.e. second cousin or closer) with age (Figure 1e), we see significant and opposing age effects for different classes of PR (Figure 1f). We note that although these trends are highly significant, the changes are subtle; for example, 23% of British Pakistani individuals aged 70-80 were inferred to be offspring of first cousins or closer, compared to 38% of those aged 15-30 (Supplementary Figure 1).

### Associations between autozygosity and common confounders

We then examined associations between F_ROH_ and phenotypes in G&H and UKB, considering two sets of individuals within each cohort: we carried out one version of the analyses using all individuals (full cohort) and one using only individuals who are inferred to be offspring of first cousin/avuncular unions and who have F_ROH_ < 0.18 (highly consanguineous cohort). (The cutoff of F_ROH_<0.18 was chosen as it is the midpoint between the expected F_ROH_ for individuals having avuncular versus sibling parents.) The motivation for this was that we suspected that social and environmental correlates of consanguinity may confound associations between phenotypes and F_ROH_ within the full cohort, i.e. highly consanguineous individuals might have systematically different cultural, social, or environmental exposures to those whose parents are unrelated. If we restrict to individuals whose parents had the same degree of PR and control for population structure, variance in F_ROH_ is attributable to stochastic recombination events and Mendelian segregation (Figure 2), thus mitigating associations between F_ROH_ and environmental confounders. We excluded a small number of individuals with F_ROH_ > 0.18 whose parents may be first-degree relatives, since such unions might be associated with extreme environmental confounders.

**Figure 2.**
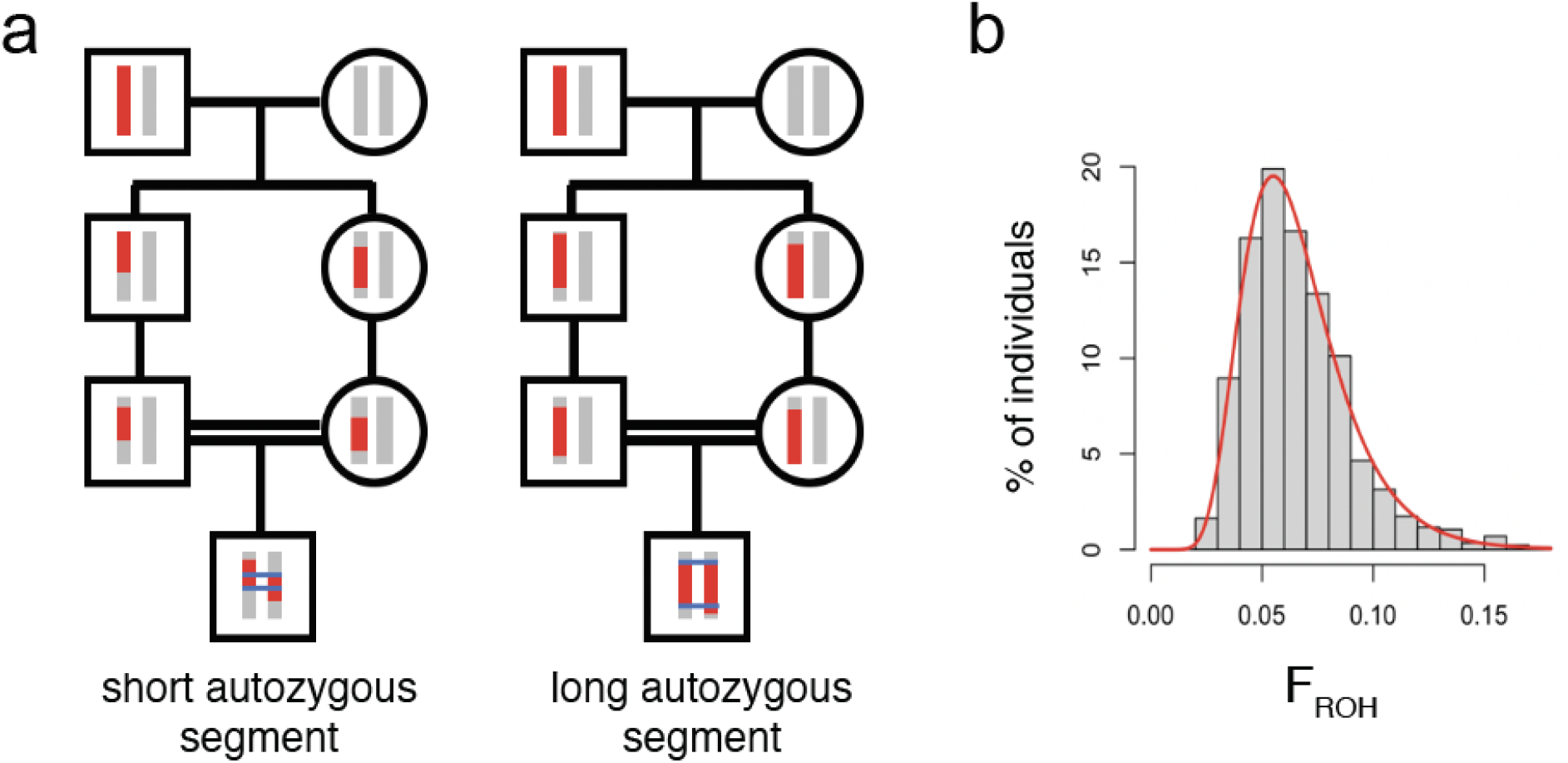
Variability in autozygosity due to stochastic recombination and Mendelian segregation events among individuals with parents who are first cousins. (a) Figure illustrating, using just one chromosome, how autozygosity can vary substantially between individuals who are offspring of first cousins. Two offspring of independent first cousin unions have inherited different ROHs of different lengths on one chromosome due to stochastic recombination and Mendelian segregation events. This leads to the variation in genome-wide F_ROH_ shown in panel (b) for G&H individuals inferred to have parents who are first cousins. The red line in (b) indicates the best fit of a lognormal distribution, which was used for power calculations.

To test the robustness of this approach, we considered five traits/exposures which may confound associations with F_ROH_ in UKBEUR and UKBSAS: self-reported religiosity, ever smoked tobacco, ever drank alcohol, socioeconomic status as measured by the Townsend Deprivation Index (SES), and having attended university. Clark et al. previously showed that F_ROH_ negatively correlated with educational attainment (EA) and alcohol and tobacco use^8^. We find that in the full cohort, F_ROH_ is significantly associated with all five traits assessed in UKBEUR and UKBSAS (Figure 3). However, in the highly consanguineous cohorts we find no significant associations. Using power calculations^28^, we find that the power to detect significant associations in the highly consanguineous cohorts using the OR estimated from the full cohorts ranges from 0.72 to >.99 with a median of 0.86, suggesting the widespread attenuation observed was unlikely to be due to the reduction in sample size when restricting to the highly consanguineous cohorts.

**Figure 3.**
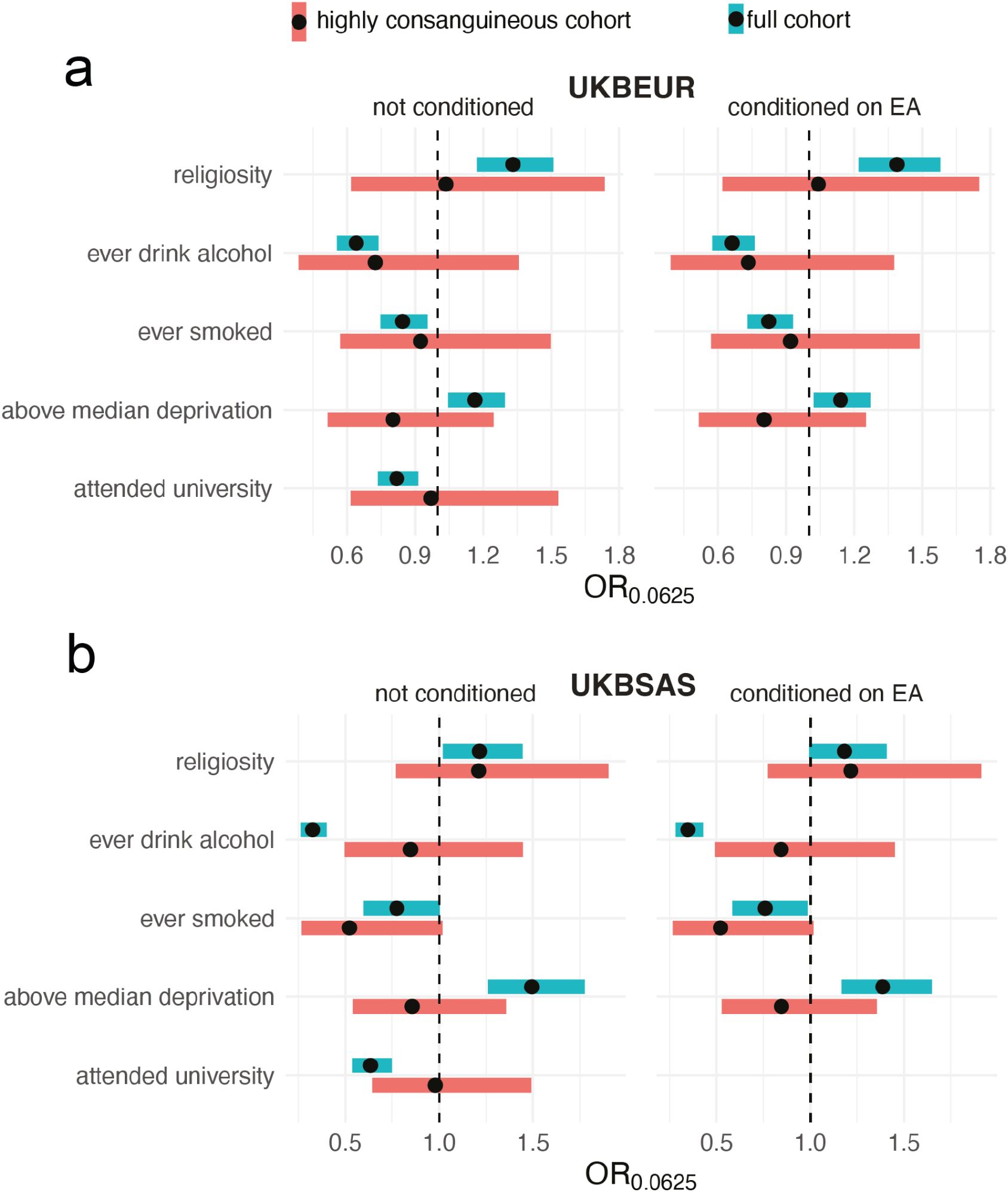
Associations between F_ROH_ and potential confounders with and without conditioning on educational attainment in (a) UKBEUR and (b) UKBSAS. Forest plot showing F_ROH_ odds ratio. OR is calculated for F_ROH_ value of 0.0625 (expected F_ROH_ for first cousin PR). Bands indicate 95% confidence intervals adjusted for multiple testing (p<.05/9).

As has been done in previous work to attempt to control for confounding^8,9,29^, we then repeated these analyses controlling for educational attainment (EA; specifically, number of years in education). This made minimal difference to our results (Figure 3, right), showing that conditioning on EA does not attenuate associations with the potential confounders we considered.

### Associations between autozygosity and disease

Having demonstrated that focusing on highly consanguineous individuals attenuates confounding with risk factors for ill health (Figure 3), we then assessed associations between F_ROH_ and diseases in this subset of individuals, meta-analysing G&H and UKB. To define the disease phenotypes, we used the first-occurrence three letter ICD-10 codes in UKB and generated phenotypes in G&H by mapping diagnostic codes from primary and secondary care EHRs using the methods defined in UKB (Methods, Supplementary Methods). We considered the sixty-one diseases with at least a 5% case prevalence in the G&H highly consanguineous cohort, since this was the largest sample (N=4,034 versus N=977 and N=754 for UKBEUR and UKBSAS respectively).

After 5% FDR correction, we find twelve associations, with four associations passing Bonferroni correction (p<0.05/61) in the meta-analysis of the highly consanguineous cohorts (Figure 4a, Supplementary Table 1). The disorders span several organ systems including metabolic, psychiatric, ear, eye, immune, and respiratory disorders. We assessed whether the effect of F_ROH_ varied linearly with respect to the log-odds, using binned F_ROH_ values, to ensure model assumptions were met. We find that the increase in log-odds for the significant traits consistently appears to be approximately linear (Supplementary Figure 2), suggesting the associations are not driven by extreme F_ROH_ values.

**Figure 4.**
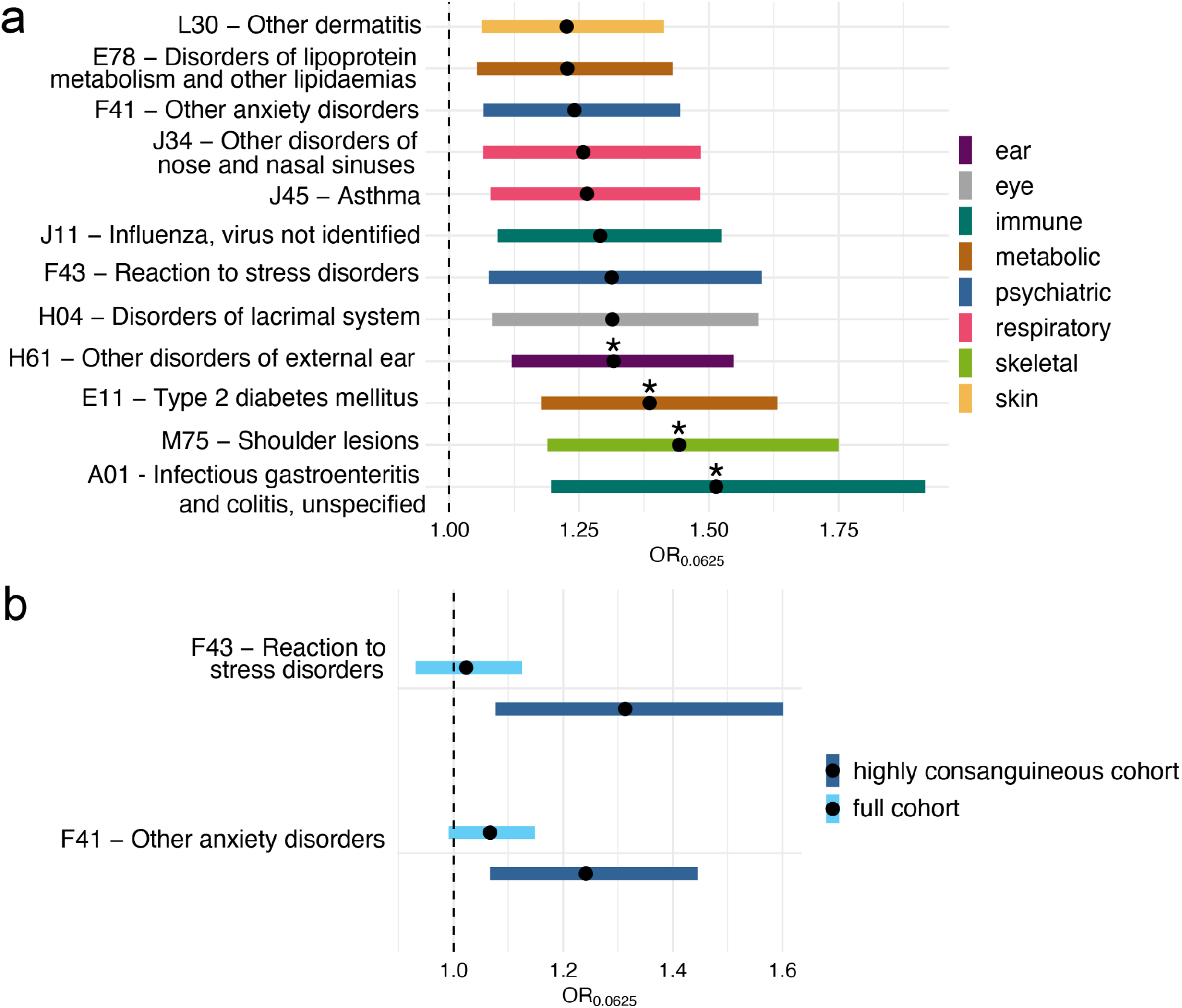
Associations between F_ROH_ and disorders significant after 5% FDR correction in the meta-analysis of highly consanguineous cohorts from G&H and UKB. (a) shows all significant disorders, and (b) highlights two psychiatric disorders that showed significant associations in the meta-analysis of highly consanguineous cohorts but not of full cohorts. Forest plot showing F_ROH_ odds ratio (OR). OR is calculated for F_ROH_ value of 0.0625 (expected F_ROH_ for first cousin PR). Bands indicate 95% confidence intervals, asterisks indicate traits that pass Bonferroni correction(p<0.05/61) and colours indicate disorder categories.

When conducting the same analysis in the full cohort, thirty traits were significant after FDR correction and thirteen passed Bonferroni correction (Supplementary Table 2). The highly consanguineous and full cohort analyses share ten significant associations at FDR<5%, with the two psychiatric traits being unique to the former (Figure 4b). We observe an inflation in the p-values for Cochran’s Q test for heterogeneity in the meta-analysis of the full cohorts and none for the highly consanguineous cohorts (Supplementary Figure 3), suggesting the effect size estimates are more consistent across the different highly consanguineous cohorts.

### Between-sibling analysis of F_ROH_-phenotype associations in 23andMe

To attempt to replicate findings, we conducted a between-sibling analysis in the 23andMe cohort using self-reported phenotypes (n=42,218-545,806, median 478,590; Supplementary Table 3). This complementary approach exploits variation in F_ROH_ within nuclear families, which eliminates confounding due to population structure ^8,30,31^. Confirming the results in Figure 3, we found no significant association (p>0.15) between F_ROH_ and having ever used tobacco or reporting being ‘at all religious’. We then considered fourteen disease phenotypes that match or are similar to the three-digit ICD10 codes that passed FDR<5% in the meta-analysis of either the highly consanguineous and/or full cohorts from G&H+UKB (Supplementary Table 3). The seven diseases that were significant in the G&H+UKB highly consanguineous cohorts showed convincing evidence of replication: all had concordant directions of effect size, significantly more than expected by chance (p=0.008, one-sided binomial test), and two were experiment-wide significant [post-traumatic stress disorder, included within ICD10 chapter F43 (OR=1.96, p=0.00082) and type 2 diabetes (OR=1.57, p =0.00395)]. In contrast, of the seven diseases that were only significant in the G&H+UKB full cohorts, five had discordant directions of effect in 23andMe and none passed experiment-wide significance. Importantly, PTSD, the disorder with the most significant F_ROH_ association in the replication analysis, was only significant in the analysis of the highly consanguineous cohorts in G&H and UKB (Figure 4b).

### Population attributable risk of autozygosity to T2D and asthma

British South Asians have more than twice the rate of T2D compared to White British Europeans^12,32^, as well as a higher rate of asthma hospitalizations and death^12,32^. Given the detected associations between autozygosity and these diseases, we estimated the fraction of the incidence of these disorders that may be attributable to the rate of consanguinity in each population. To do so, we calculated the percent population attributable risk (i.e. percent of cases in the population attributable to autozygosity) for the two diseases, using the odds ratio estimates for F_ROH_ from the G&H+UKB meta-analysis of the highly consanguineous cohorts and the rate of consanguinity estimated in UKBEUR individuals and in G&H British Bangladeshi and Pakistani individuals (see Methods). Since conversion of the odds ratio estimate requires an estimate of the prevalence of the disorders in nonconsanguineous individuals, which is not available, we varied the assumed prevalence from 5% to 15% for each disorder, as that should reasonably capture the true prevalence^33,34^.

Assuming a 5% prevalence of disease in nonconsanguineous individuals, we estimated that 10.1% (5.2%-15.9%, 95% CI) of the prevalence of T2D in G&H British Pakistanis is attributable to autozygosity resulting from consanguinity (Figures 5 and S4a-d). This is independent of the environmental/cultural correlates of consanguinity that may influence risk of the disorder. The rate was estimated at 2.6% (1.2%-4.6%) in G&H British Bangladeshis, and at <1% in UKBEUR. Likewise, we estimated 7.4% (2.5%-12.5%) of asthma cases in G&H British Pakistanis are attributable to autozygosity, 2.4% (0.9%-4.2%) in G&H British Bangladeshis, and <1% in UKBEUR. The estimates increase slightly when assuming a prevalence of 15%. Thus, we conclude a substantial proportion of the increased incidence of T2D in British Pakistanis is due to autozygosity resulting from consanguinity.

**Figure 5.**
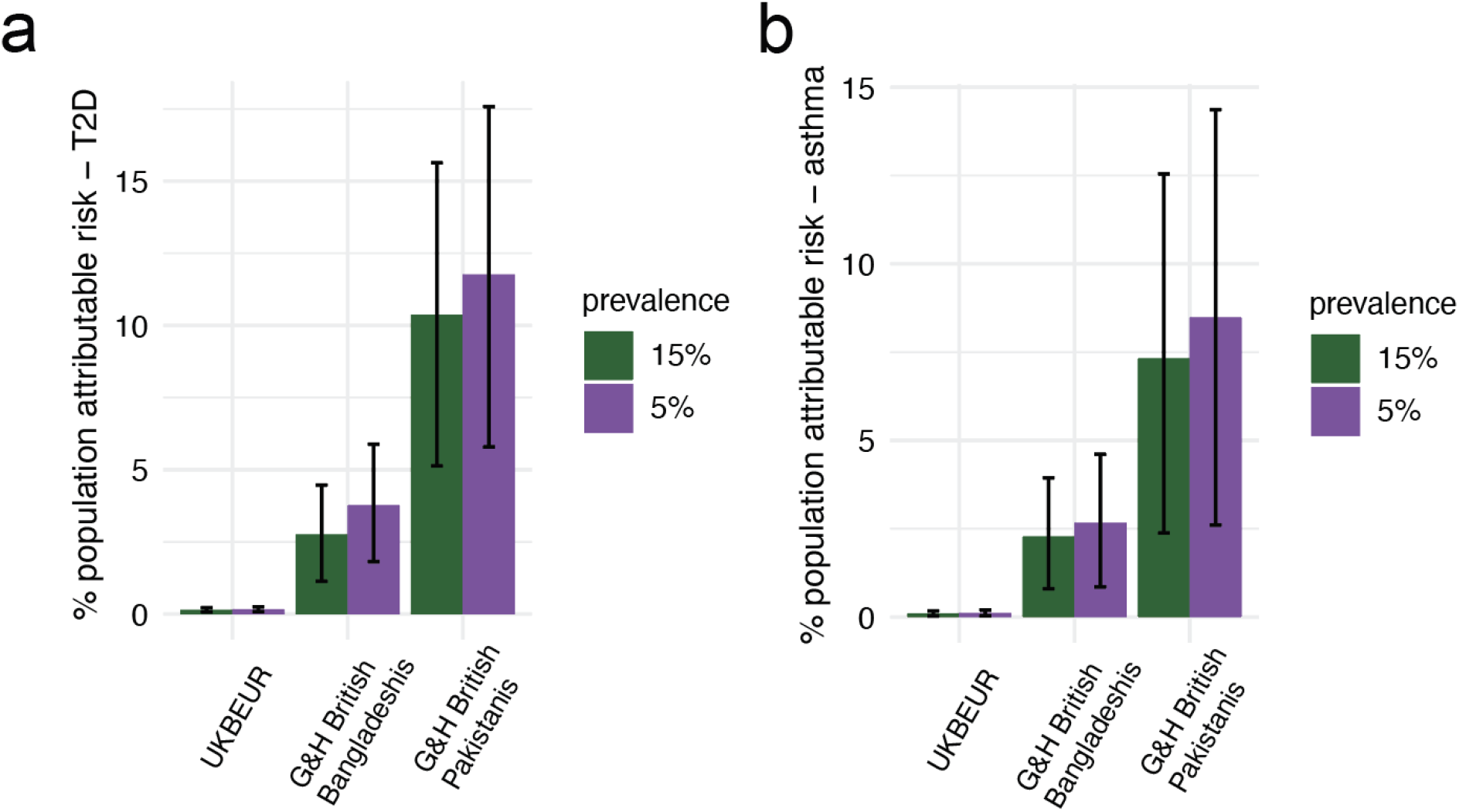
Percent population attributable risk of F_ROH_ on (a) T2D and (b) asthma estimated for UKBEUR and G&H British Bangladeshis and Pakistanis, assuming underlying prevalence estimates of disease in the nonconsanguineous population equal to 5% or 15%. Error bars indicate 95% confidence intervals.

As a point of comparison for T2D, we considered the population attributable risk due to having a high polygenic risk score (PRS) for the disease. We considered the T2D PRS developed in Mars et al.^35^ which showed similar predictive accuracy in European and British South Asian cohorts they studied (OR for 1SD of the PRS ∼1.65 in both populations). Supplementary Figure 4e shows that in G&H British Pakistanis and British Bangladeshis, the increase in T2D prevalence due to autozygosity is similar to that due to individuals being in the top 5-18% and 1-3% of polygenic risk, respectively.

### Impact of genetic architecture on F_ROH_ associations with binary traits

Associations between F_ROH_ and traits can be induced by several underlying genetic architectures. A commonly described hypothesis is that F_ROH_ increases the risk of inheriting deleterious recessive variants, thereby increasing genetic predisposition towards disease. An alternative (but not mutually exclusive) explanation is that autozygosity increases the additive genetic variance of a trait in the population (specifically by a factor of 1+*F*, where *F* is the average “inbreeding coefficient” in the population^16^, also see Supplementary Note). Thus, under a liability threshold model for a binary trait, individuals with high values for F_ROH_ are more likely to cross the liability threshold even in the absence of non-additive effects, inducing an association between F_ROH_ and the trait (Supplementary Figure 5).

To assess the degree to which the increased additive variance could induce associations between F_ROH_ and binary traits, we simulated binary traits with an additive polygenic genetic architecture and varying heritabilities, then estimated the power we would have to detect significant associations between F_ROH_ and the simulated traits in our current study, considering the sample size and F_ROH_ distribution in the highly consanguineous cohorts. We find that purely additive, polygenic traits with heritabilities similar to those of the most heritable traits we consider (e.g. T2D with an estimated narrow-sense h^2^ of 20%-30%^36^) would be very underpowered to show significant associations with F_ROH_ in our study (Supplementary Figure 6a). In the unrealistic case of F_ROH_ values in the population being uniformly distributed from 0 to 1, there would still be very little power to detect associations at our current sample size (Supplementary Figure 6b). We conclude that the associations we observe are unlikely to reflect a solely additive genetic architecture and hence highlight the possibility of widespread non-additive effects on diseases across the phenotypic spectrum.

## Discussion

We introduce a novel approach to reduce confounding in studies assessing trait associations with autozygosity by restricting analyses to highly consanguineous individuals. We find compelling evidence that autozygosity impacts several common diseases spanning multiple organ systems, notably type 2 diabetes and PTSD. Simulations indicate that the associations most likely stem from non-additive genetic effects, and we calculate population attributable fractions to show that these effects cumulatively contribute substantially to disease incidence in populations with high rates of consanguinity.

In concordance with previous studies^1,37,38^, we find that British Bangladeshi and Pakistani individuals practise consanguinity at higher rates than British individuals with European ancestries. Our results from G&H show that younger British Pakistanis are more likely to have parents inferred to be first cousins, while overall consanguinity (i.e. second cousin or closer) is decreasing in younger British Bangladeshis. The fact that we see opposite patterns in the two groups suggests that this is not due to the impact of autozygosity on health which could lead to ascertainment biases. We cannot be sure whether the patterns we observe are due to changing patterns of unions *within the UK* across time or temporal changes in migration rates from Pakistan/Bangladesh to the UK which affected trans-national marriage/union patterns^39^. Recent work in large biobank settings has shown that overall rates of consanguinity are decreasing in large cohorts from the United States (All of Us and Million Veterans Program) and increasing in UKB South Asians^27^. Our analysis suggests that examining these trends at the level of a whole country or broad ancestry group may obscure fine-scale differences. Also, considering only changes in mean F_ROH_ may obscure changes in rates of different types of consanguinity (Figure 1 and Supplementary Figure 1). These results highlight important trends for clinical settings, as autozygosity increases the risk of recessive Mendelian diseases^40^ and, as we show here, several common, complex disorders.

Before we assessed association between F_ROH_ and disease, we investigated associations between F_ROH_ and common confounders that are associated with disease risk, including socioeconomic, behavioural, and cultural traits in the UKB cohorts. When considering all individuals, we found significant associations between F_ROH_ and university attendance, deprivation, religiosity and alcohol/tobacco use. All of the associations were attenuated with our approach of restricting to the highly consanguineous cohort, suggesting they were due at least in part to confounding. Consistent with this, religiosity and tobacco use were likewise not significant in the 23andMe between sibling-analysis. We found that conditioning on EA, a sensitivity analysis common in the autozygosity literature^8,9,29^, did not attenuate the associations between F_ROH_ and the potential confounders assessed (Figure 3). These analyses illustrate the need to carefully assess whether the causes of F_ROH_ associations in several previous studies are indeed biological, and emphasise that they should be interpreted with caution.

Having demonstrated that restricting analyses to highly consanguineous individuals greatly attenuates confounding, we investigated associations between F_ROH_ and clinical phenotypes extracted from EHRs within this group. We found associations between F_ROH_ and twelve diseases classified by three-digit ICD10 codes, including T2D, asthma, and two psychiatric disorders (“F43 - reaction to severe stress disorders”, which includes PTSD, and “F41 - other anxiety disorders”). We also replicated T2D and PTSD at experiment-wide significance in the 23andMe between-sibling analysis. We saw several additional associations in G&H+UKB, including shoulder lesions, which include adhesive capsulitis, a common comorbidity of T2D^41^. Additionally, it has been shown that PTSD symptoms and diagnosis are associated with increased risk for T2D^42^. As cohorts with highly consanguineous individuals grow and non-additive loci are discovered for these disorders, it may be possible to disentangle the potential causal paths operating between these associations.

When analysing the full cohort from G&H+UKB, we found multiple additional associations. However, when attempting to replicate seven of these via between-sibling analysis in 23andMe, none passed experiment-wide significance and five had discordant directions of effect size, indicating they were likely spurious. Interestingly, the analysis of the full G&H+UKB cohorts gave nonsignificant results for the two psychiatric disorders identified in the highly consanguineous analysis (Figure 4b). This result suggests that environmental/cultural factors correlated with consanguinity, and therefore F_ROH_, in these cohorts are either truly protective against these disorders, and/or that consanguineous individuals are less likely to seek medical assistance for them. Thus, our approach not only addresses spurious associations between F_ROH_ and diseases, but also prevents masking that is potentially due to consanguinity-related differences in disease ascertainment in EHRs.

Our paper has several limitations. Our approach assumes that within the highly consanguineous subset of the cohort, the degree of autozygosity is not correlated with environmental factors that influence disease risk, which we cannot totally rule out. However, our results in Figure 3 suggest that there is no remaining association with some obvious potential confounders. Another limitation of the paper is that, after multiple testing correction, we only replicated two of the seven diseases tested in the between-sibling analysis from 23andMe. This is likely due to the more limited power of this approach, but results for these other diseases should be treated with caution unless replicated in future studies.

We showed that the risk towards T2D and asthma incurred by autozygosity may contribute substantially to the incidence of these diseases in British Pakistanis, and to a lesser degree in British Bangladeshis. Our estimates of population attributable risk (PAR) assume that G&H is representative of the broader British Pakistani and Bangladeshi communities in the UK, though we note the majority of the current G&H cohort is from London. Our recent work in a cohort collected in Bradford, a city in the north of England with a substantial British Pakistani population, reported higher rates of consanguinity than found here, with 44% of British Pakistanis inferred to have parents who are first cousins or closer^2^ (compared to 33% in the current study), suggesting we are potentially underestimating the true PAR. For T2D, we found that the rate of consanguinity in British Pakistanis increases the prevalence in the population approximately equivalently to individuals being in the top decile of common variant risk measured in a previous study^35^. Importantly, we note that our estimates for the PAR due to autozygosity have large standard errors (the confidence intervals for T2D for British Pakistanis span between 5.2% and 17.5%, depending on assumed prevalence), and that other risk factors for T2D have a far higher PAR than autozygosity. One study estimated the PAR for having BMI > 25 kg/m^2^ is >60% in the Americas, with little fluctuation between regions^43^. In a separate study of a cohort based in Rotterdam, the PAR for BMI > 25 kg/m^2^ was 51%, and 71% for all modifiable risk factors assessed in their study (high BMI and waist circumference, current smoking, and high C-reactive protein)^44^. Thus, while the impact of autozygosity resulting from consanguinity on T2D risk is significant, its impact is less than other, modifiable risk factors. Furthermore, the health risks incurred by consanguinity need to be weighed against potential social and economic benefits for communities.

Via simulations, we show that the associations we detected are unlikely to be due to autozygosity increasing additive variance for genetic risk of binary traits, suggesting wide-spread non-additive effects. In the few studies that have looked, recessive-acting rare and common variants have been found to be associated with multiple common diseases including T2D^45–47^. However, it has been previously shown that dominance heritability at common variants is negligible^48,49^, suggesting the observed F_ROH_ associations likely stem from non-additive effects at low allele frequency variants and/or epistasis. Assuming an outbred population, detecting recessive effects requires much larger sample sizes than for additive loci, since only *np*^*2*^ individuals have informative alternative genotypes (where *n* is sample size and *p* is the effect allele frequency) versus *n*(*p*(1-*p*)+*p*^*2*^))=*np* under an additive model. This issue is especially exacerbated at rare variants due to the quadratic scaling, but is reduced in consanguineous cohorts where the number of informative alternative genotypes for recessive loci is *n*((1-mean(*F*))*p*^*2*^ + mean(*F)p*). Thus, large sequenced cohorts from populations with high levels of consanguinity will be necessary to fully characterise the nature of non-additive genetic effects across the allele frequency spectrum for polygenic traits.

In conclusion, we have described patterns of consanguinity in two large UK cohorts and proposed a novel approach to control for social and environmental confounding in autozygosity association studies. We found multiple significant associations between autozygosity and common diseases which we contend are unlikely to be confounded. Our findings suggest that previous results in the field should be revisited, as they may have been driven by uncontrolled confounders. Furthermore, our results indicate that autozygosity may be an important contributing factor to the increased incidence of T2D in British Pakistanis as well as in other worldwide populations with high rates of consanguinity. Our work motivates the incorporation of genome-wide autozygosity into predictions of genetic risk, as well as a search for individual non-additive-acting variants and genes influencing disease risk across the phenotypic spectrum.

## Supporting information

Frequently Asked Questions

## Data Availability

G&H data are available for analysis within a secure Trusted Research Environment) upon application to the G&H executive, as described here https://www.genesandhealth.org/research/scientists-using-genes-health-scientific-research. UK Biobank data are also available upon application (https://www.ukbiobank.ac.uk/enable-your-research/apply-for-access).

## Acknowledgements

We thank Loic Yengo, Richard Durbin, Peter Visscher, John Perry, Nicole Soranzo and Matt Hurles for useful discussions, and Muhammad Forhad and Naheed Choudhry from the G&H Community Advisory Board for their help on the Frequently Asked Questions document.

Authors from the Wellcome Sanger Institute are supported by Wellcome grant 220540/Z/20/A, ‘Wellcome Sanger Institute Quinquennial Review 2021-2026’. D.M. is supported by a Gates Cambridge Scholarship (OPP1144).

Genes & Health is/has recently been core-funded by Wellcome (WT102627, WT210561), the Medical Research Council (UK) (M009017, MR/X009777/1), Higher Education Funding Council for England Catalyst, Barts Charity (845/1796), Health Data Research UK (for London substantive site), and research delivery support from the NHS National Institute for Health Research Clinical Research Network (North Thames). Genes & Health is/has recently been funded by Alnylam Pharmaceuticals, Genomics PLC; and a Life Sciences Industry Consortium of Astra Zeneca PLC, Bristol-Myers Squibb Company, GlaxoSmithKline Research and Development Limited, Maze Therapeutics Inc, Merck Sharp & Dohme LLC, Novo Nordisk A/S, Pfizer Inc, Takeda Development Centre Americas Inc. Additional funding for this work was awarded by the Medical Research Council (MR/S027297/1).

We thank Social Action for Health, Centre of The Cell, members of our Community Advisory Group, and staff who have recruited and collected data from volunteers. We thank the NIHR National Biosample Centre (UK Biocentre), the Social Genetic & Developmental Psychiatry Centre (King’s College London), Wellcome Sanger Institute, and Broad Institute for sample processing, genotyping, sequencing and variant annotation. We thank: Barts Health NHS Trust, NHS Clinical Commissioning Groups (City and Hackney, Waltham Forest, Tower Hamlets, Newham, Redbridge, Havering, Barking and Dagenham), East London NHS Foundation Trust, Bradford Teaching Hospitals NHS Foundation Trust, Public Health England (especially David Wyllie), Discovery Data Service/Endeavour Health Charitable Trust (especially David Stables), NHS Digital - for GDPR-compliant data sharing backed by individual written informed consent.

Most of all we thank all of the volunteers participating in Genes & Health and UK Biobank.

Current <u>Genes & Health Research Team</u> (in alphabetical order by surname): Shaheen Akhtar, Mohammad Anwar, Elena Arciero, Omar Asgar, Samina Ashraf, Gerome Breen, Raymond Chung, Charles J Curtis, Shabana Chaudhary, Maharun Chowdhury, Grainne Colligan, Panos Deloukas, Ceri Durham, Faiza Durrani, Fabiola Eto, Sarah Finer, Ana Angel Garcia, Chris Griffiths, Joanne Harvey, Teng Heng, Qin Qin Huang, Matt Hurles, Karen A Hunt, Shapna Hussain, Kamrul Islam, Ben Jacobs, Ahsan Khan, Amara Khan, Cath Lavery, Sang Hyuck Lee, Robin Lerner, Daniel MacArthur, Daniel Malawsky, Hilary Martin, Dan Mason, Mohammed Bodrul Mazid, John McDermott, Sanam McSweeney, Shefa Miah, Sabrina Munir, Bill Newman, Elizabeth Owor, Asma Qureshi, Samiha Rahman, Nishat Safa, John Solly, Farah Tahmasebi, Richard C Trembath, Karen Tricker, Nasir Uddin, David A van Heel, Caroline Winckley, John Wright.

This research has been conducted using the UK Biobank Resource, a major biomedical database, under Application Number 44165.

We would like to thank the research participants and employees of 23andMe for making this work possible. The following members of the 23andMe Research Team contributed to this study: Stella Aslibekyan, Adam Auton, Elizabeth Babalola, Robert K. Bell, Jessica Bielenberg, Katarzyna Bryc, Emily Bullis, Daniella Coker, Gabriel Cuellar Partida, Devika Dhamija, Sayantan Das, Sarah L. Elson, Nicholas Eriksson, Teresa Filshtein, Alison Fitch, Kipper Fletez-Brant, Pierre Fontanillas, Will Freyman, Julie M. Granka, Karl Heilbron, Alejandro Hernandez, Barry Hicks, David A. Hinds, Ethan M. Jewett, Yunxuan Jiang, Katelyn Kukar, Alan Kwong, Keng-Han Lin, Bianca A. Llamas, Maya Lowe, Jey C. McCreight, Matthew H. McIntyre, Steven J. Micheletti, Meghan E. Moreno, Priyanka Nandakumar, Dominique T. Nguyen, Elizabeth S. Noblin, Jared O’Connell, Aaron A. Petrakovitz, G. David Poznik, Alexandra Reynoso, Morgan Schumacher, Anjali J. Shastri, Janie F. Shelton, Jingchunzi Shi, Suyash Shringarpure, Qiaojuan Jane Su, Susana A. Tat, Christophe Toukam Tchakouté, Vinh Tran, Joyce Y. Tung, Xin Wang, Wei Wang, Catherine H. Weldon, Peter Wilton, Corinna D. Wong.

## Author contributions

D.S.M. helped conceive the project, conducted the analyses and wrote the first draft of the manuscript. E.v.W. helped conceive the project and conducted quality control, ROH calling and analyses in the UKB data. B.J. processed the G&H phenotype data. Q.Q. T.H.H. and D.S.M. conducted quality control on the G&H genotype data. A.H.S, S.R., S.M.S. and A.K. assisted with writing the manuscript and the FAQ document. M.U.M. and D.P. helped supervise the preparation of the UKB data. R.M., D.v.H. and S.F. helped supervise the G&H work, and S.F. and D.v.H. supervised the collection of the G&H data. J.O. helped conceive the project and contributed intellectually to the analyses. H.C.M. conceived and directed the project and helped draft the initial manuscript. All authors commented on the manuscript.

## Disclosures

J.O. and members of the 23andMe Research Team are employed by and hold stock or stock options in 23andMe, Inc. Other authors report no disclosures.

## Methods

### G&H and UK Biobank cohorts and genotype data preparation

We used the 2021 July data release of the G&H data, which contained 46,132 individuals genotyped on the Illumina Global Screening Array v3EAMD (GRCh38). The G&H cohort was recruited across several sites in East London, Luton, Manchester, and Bradford, including community settings (e.g. mosques, shopping centres, libraries) and primary care clinics^26^. Fifty six percent of individuals were recruited in primary care settings, 5% were recruited in hospitals, and the remaining were recruited in community settings. We first removed 1,736 individuals with call rate less than 99.2% and SNPs with MAF < 1%, leaving 355,862 SNPs. To ensure we did not lose SNPs that have high quality but that fail Hardy-Weinberg Equilibrium due to high rates of consanguinity and strong population structure in British Pakistani individuals, we removed 726 SNPs that failed Hardy-Weinberg Equilibrium p-value < 1×10^−6^ in British Bangladeshi individuals alone, as done in Huang et al.^50^ This left 355,136 SNPs.

Genotyping and processing for the UK Biobank cohort were done centrally by the UKB group^51^. Two customized Affymetrix genotyping arrays were used, the UK Biobank Axiom array (n=438,692) and the UK BiLEVE Axiom array (n=50,520), which covered 812,428 SNPs with 95% overlap between the arrays. Quality control consisted of excluding individuals with >3% missingness, inconsistent sex, sex aneuploidy, excess heterogeneity, or withdrawn consent.

In both cohorts, we estimated the relatedness between individuals using PropIBD from KING^52^ removed one from each pair of related individuals inferred to be 3^rd^ relatives or closer. To remove related individuals while maximising the sample size, we ranked individuals by their number of relatives, then removed the individual with the highest number of relatives until no relatives remained.

### Ancestry definition

In G&H, we inferred genetic ancestry by merging the data with reference sequences of unrelated individuals (determined using KING as described above) from the 1000 Genome Project^53^ and Central and South Asian individuals from the Human Genome Diversity Project^54^. We first excluded palindromic variants and multiallelic sites from both datasets. Then, we merged the external reference data and G&H by matching positions and alleles of the common SNPs that passed QC in G&H, and kept variants found in both datasets, which left 349,632 SNPs. A further 1285 variants were excluded due to AF discrepancies between G&H and South Asian reference individuals (>4 standard deviations from the mean residual of -log10 frequency bins, and Fisher’s exact test p<1×10^−5^), resulting in 348,347 variants. PLINK 1.9 LD pruning was performed with a window size of 1000kb, step size 50 and LD r^2^ cutoff of 0.1, then long LD regions^55^ were excluded, resulting in 104,552 variants. We used PLINK 1.9 to calculate principal components (PCs). To remove individuals with non-South Asian ancestry, we first calculated PCs for the 3,433 reference individuals, projecting the G&H samples into the reference PC space. We calculated UMAP coordinates using the umap R package. We found that the UMAP with 7 PCs was optimal to separate the reference individuals into their assigned superpopulations. 44,320 out of 44,396 G&H individuals were inferred to be South Asians at this stage. To identify British Bangladeshi and Pakistani individuals in G&H, we then performed a second PC analysis on the unrelated G&H individuals, projecting the related G&H individuals into the PC space defined by the unrelateds. The UMAP with 4 PCs identified distinct Pakistani and Bangladeshi clusters (defined based on the self-reported ancestries), and this was used to classify individuals as genetically Pakistani or Bangladeshi (leaving 44,190 individuals in the final dataset).

UKB participants were divided into five continental groups, defined by projecting UKB individuals into the 1000 Genomes PCA space. Individuals were then grouped into their closest ancestral population, based on the Mahalanobis distance between their projected principal component score and the average score of each ancestral sample. Individuals with a Mahalanobis distance that deviated from each population average at >6 SD were excluded. 387,531 individuals of European descent and 9,653 individuals of South Asian descent remained after quality control.

### ROH calling

For ROH calling in G&H, we filtered out SNPs with minor allele frequency <5% and used PLINK 1.9 to call ROHs on the filtered SNPs using the following parameters, following Clark et al.^8^: -homozyg-window-snp 50 --homozyg-snp 50 --homozyg-kb 1500 --homozyg-gap 1000, --homozyg-density 50 --homozyg-window-missing 5 --homozyg-window-het 1. In UKB we followed the same procedure, but before ROH calling we removed variants that had Hardy-Weinberg p<1×10^−6^ in the relevant ancestry group.

We calculated F_ROH_ by summing up the total length of all autosomal ROHs previously calculated (in base pairs) and dividing by 2.7 billion (the approximate length of the autosomal genome), following Clark et al.^8^.

### Consanguinity inference

We developed a method to infer parental relatedness which has been fully described in ^2^. Briefly, unrelated individuals were randomly chosen from the actual dataset, phased using Eagle v2.4.1^53^, and consanguineous pedigrees are simulated using custom R code available at https://github.com/malawsky/consanguinity_simulation; specifically we included unions between individuals who are siblings, avuncular (including multiple generations), first cousins (including multiple generations), first cousins once removed, and second cousins, as well as between unrelated individuals. We then applied the same ROH calling procedures described above to the simulated offspring. For each simulated individual, we then calculated fifteen statistics for the purposes of classification using the neural net classifier: the total length of the ten longest ROHs (in cM), and the frequency of ROHs ranging from 10 to 150 cM binned into 14 intervals of 10 cM. Using these statistics, we trained a neural net classifier implemented in the R package *nnet* to assign simulated individuals to a given consanguineous pedigree by repeating this procedure 10 times, summing up the probabilities for each possible PR category, and choosing the one with the highest probability per individual. We then calculated the same statistics on the true samples and used the trained neural net classifier to infer the degree of PR. For most of our analyses, we group together people whose parents were inferred to be second cousins with first cousins once removed, and people whose parents were inferred to be first cousins for one/two/three generations, because of the low accuracy in differentiating between the finer-grained classifications.

### Analysis of consanguinity patterns in G&H and UK Biobank

In G&H, individuals were asked about their parental relatedness at recruitment (“Were your parents related by blood? (not just by marriage)”) with the options of “Yes”, “No”, and “Don’t know”. If the individual answered “Yes”, they were asked a follow-up question of “If Yes, how were your parents related?” with the options of “First Cousins”, “Don’t Know’’, and “Other related by blood”. Figure 1b,c shows the inferred degree of parental relatedness for individuals split by self-reported parental relatedness.

We used linear regression to regress F_ROH_ on age G&H British Pakistanis, G&H British Bangladeshis, UKBEUR and UKBSAS, controlling for sex and 20 PCs. To test if overall consanguinity changed over time, we made a binary variable indicating parental relatedness (1 if inferred to have parents that are second cousins or closer, 0 otherwise) and regressed that on age, sex, and 20 PCs using a logistic regression. To test for more subtle changes in consanguinity patterns over time, we made a categorical variable indicating each of the three main inferred parental relatedness categories (first cousins or closer, second cousins/first cousins once removed, or unrelated), and regressed it on age, sex, and 20 PCs using a multinomial logistic regression with the nnet R package^56^.

### Association between autozygosity and traits

#### Phenotypic data harmonisation and preparation for G&H and UK Biobank

The G&H EHR data consisted of SNOMED codes from primary care data for 34,712 of the participants (i.e. those registered with a GP in inner London, outer London, and Bradford), ICD-10 codes from secondary care data for 17,132 individuals (i.e. those who had attended the Barts Health or Bradford University Hospitals NHS trusts), and ICD-10 codes from national Hospital Episode Statistics available on all participants. There were twelve participants with no ICD-10 codes, and we removed these individuals from the analyses since it was possible that they had recently moved to the UK so may be missing any EHR data for that reason. After removal of relatives, 23,978 individuals were retained. We translated SNOMED codes in primary care data to ICD-10 codes using the Interactive Map-Assisted Generation of ICD-10 Codes algorithm (using only codes with strict 1:1 mapping, as also done by UK Biobank)^57^. See Supplementary Methods for additional details. Since we suspected that coding practices might be different in different areas, and since missing EHR data could otherwise affect our results, in the analyses described below we included indicator variables to account for:

- Whether a G&H individual had primary care data from an inner London borough, outer London borough, and/or Bradford (3 binary variables)
- Whether a G&H individual had at least one secondary care code from Barts Health or Bradford University Hospitals NHS trust (2 binary variables)

To define disease phenotypes in UKB (Figure 4, Supplementary Tables 1 and 2), we used the ‘first-occurence’ ICD10 codes (field 1712). The UKB phenotypes used in Figure 3 were as follows:

- Religiosity (field 100328) indicates whether an individual reported attending a religious group at least once a week.
- Townsend deprivation index (field 189) was used as a proxy for socioeconomic status.
- Educational attainment (field 6138) was binarised into ‘having attended university’ or not when used as an outcome phenotype (for easy of comparison with the other phenotypes in the figure), but when used as a covariate (right hand side of Figure 3), we converted it to years in education as done previously^58^.
- ‘Ever drinking alcohol’ was obtained from field 1558.
- ‘Ever smoked’ was obtained from field 20160.

#### Regression analyses in G&H and UK Biobank

We considered two subsets of individuals in each cohort (G&H, UKB and UKBSAS) to identify associations between F_ROH_ and phenotypes: the full cohort including all individuals and the highly consanguineous cohort consisting of individuals inferred to have parents that are first cousins. We used logistic regression in base R for binary variables.

For G&H, as covariates in the regression we included F_ROH_, sex, age, age^2^, age*sex, genetic PCs 1-20, (has primary care code from outer London primary care data), (has primary care code from inner London primary care data), (has primary care code from Bradford), (has secondary care code from Barts Health), and (has secondary care code from Bradford University Hospitals NHS trust).

In UKB, slightly different covariates were used, including array (UKB field 22000), batch (UKB field 22000), recruitment centre (UKB field 54), and whether primary care data were available (UKB field 42040).

For the meta-analysis of disease phenotypes, we used inverse variance-weighted fixed effect meta-analysis of estimates obtained from regressions in the UKBEUR and UKBSAS cohorts and in G&H.

For the log(OR) estimates by residualized F_ROH_ quintiles (Supplementary Figure 3), we regressed F_ROH_ on the other covariates and binned F_ROH_ values by quintiles. The quintiles were defined in the G&H highly consanguineous cohort as the F_ROH_ distribution in this group was very similar to that seen in the highly consanguineous individuals from UKB (Supplementary Table 4). We then regressed a given trait on the binned F_ROH_ quintiles and meta-analysed the effect size across the three cohorts. For a linear regression of the log(OR), we used an inverse variance weighted linear regression using SE estimates for each log(OR) estimate.

#### Power analyses

We used G*Power^28^ to calculate power to detect a significant effect size for logistic regression in the highly consanguineous cohorts, assuming the effect size estimates in the full cohorts. One needs to specify the frequency of the binary phenotype in the population, the expected OR, the distribution of F_ROH_, sample size, and p-value threshold. For each trait, we used the OR estimated in the full cohort analyses, the frequency of the binary phenotype in the highly consanguineous cohort, the sample size for a given highly consanguineous cohort, a p-value threshold of 0.05, and a log-normal distribution for F_ROH_ with mean -2.5 and standard deviation of 0.5, with F_ROH_ values restricted to be between 0.02-0.18 (which approximates the empirical distribution of F_ROH_ for individuals with first cousin parents; Figure 1B).

#### Between-sibling analysis in 23andMe

We conducted a between-sibling regression analysis using individuals inferred to be full biological siblings in the 23andMe cohort, including individuals from all ancestry groups since this within-family analysis is immune to population stratification. We considered 7,363,319 23andMe customers that had consented to research and had reported age, sex and at least one of the phenotypes of interest. Sibling groups were identified as cliques sharing 2249cM < IBD1 < 3373cM and 375cM < IBD2 < 2249cM^59^. We then performed relatedness pruning to avoid (for example) two generations of a pedigree being analysed as independent sibling groups. For each phenotype, only cliques containing at least two individuals with non-missing data were considered, we then greedily removed cliques with the highest number of related cliques until no clique interconnections were remaining. Two cliques were considered connected if at least one pair across the cliques shared IBD1 > 700cM. This resulted in between 20,713 and 262,433 sibling cliques containing 42,218 to 545,806 individuals depending on phenotype. ROHs were called and F_ROH_ was determined in the same way as described above for G&H and UKB.

As 23andMe does not have electronic health records, we used self-reported phenotypes as proxies to replicate our significant findings from the meta-analysis of G&H and UKB. The results and lists of equivalent phenotypes are shown in Supplementary Table 3. Using the *bife* R package^60^, we regressed the binary disease phenotype of interest on F_ROH_, adjusting for age, age^2^, sex, and sex*age as fixed effects and family membership as a random effect (i.e. family-specific intercept). For quantitative phenotypes, we regressed the phenotype on the same random and fixed effects using the *plm* R package^61^. The analysis was conducted separately for three different genotyping chips, and the results meta-analysed.

Participants provided informed consent and volunteered to participate in the research online, under a protocol approved by the external AAHRPP-accredited IRB, Ethical & Independent (E&I) Review Services. As of 2022, E&I Review Services is part of Salus IRB (https://www.versiticlinicaltrials.org/salusirb).

### Calculating population attributable risk

To calculate population attributable risk as a percentage, we used the following formula:

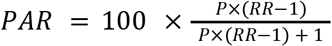

where *P* is the prevalence of the disorder in “unexposed” individuals (in our case, those with unrelated parents) and *RR* is the risk ratio for the disease. As it is not possible at present to derive robust estimates of disease prevalence excluding individuals with related parents, we varied the disease prevalence from 5% to 15% for both diseases. In practice we found that this made little difference to our estimates (Figure 5).

To convert the OR to RR, we used the following formula:

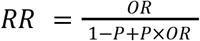

where *P*is the prevalence of the disorder in unexposed individuals and OR is the odds ratio for a given level of autozygosity on a given disorder. When estimating PAR it is necessary to discretise continuous measures, so we chose to discretise F_ROH_ into the values corresponding to the expectation for first-cousin PR (F_ROH_ = 0.0625) and second-cousin PR (F_ROH_ = 0.01562). We estimated the prevalence of these based on the estimates of the frequency of first cousin PR and of second cousin/first cousins once removed PR (Figure 1).

To calculate PAR for T2D attributable to a PRS, we used the OR estimates for a PRS developed in Mars et al.^35^ using the GWAS in ^62^, which they showed to have roughly equivalent degrees of predictive power in individuals with European versus South Asian ancestry. We used the same procedure as above, but calculated a risk ratio for individuals in twenty bins ranging from the top 1% to the top 20% of the PRS distribution, then calculated the cumulative sum of the PAR attributable to each 1% increment (Supplementary Figure 4e).

### Simulation of binary traits with strictly additive genetic architectures

We simulated the architecture of an additive binary trait by first assigning an effect size β drawn from N(0,1) for 1,000 independent causal loci. (We note that varying the parameter for the number of causal loci has no effect on our conclusions, as the genetic liability distribution for polygenic traits is normally distributed.) The allele frequency for each locus in the population was calculated by first calculating 1/β for each SNP and then linearly scaling the values to be between 0 and 0.5, to approximate model assumptions used in ^63^. (However, we note that the MAF-effect size relationship does not impact results as the additive variance and F_ROH_ relationship is not affected.) We then simulated 6,000 individuals (slightly more than the number of individuals in the combined highly consanguineous cohorts) to have F_ROH_ values either uniformly drawn from 0 to 1 or from the F_ROH_ distribution of the G&H highly consanguineous cohort (as shown in Figure 2b). For each individual, a random subset of round(1,000*F_ROH_) SNPs were assigned to be autozygous. We then simulated genotypes for each individual with the genotype in non-autozygous segments drawn from Binomial(2,*p*) and from autozygous segments from 2*Binomial(1,*p*) where *p* is the frequency of the effect allele at the locus. Genetic risk was then calculated by multiplying each individual’s genotypes with their corresponding effect sizes, summing them up, and then normalising the values across the cohort.

We then simulated a binary phenotype by drawing random values from Binomial(1,*p*_*d*_) where *p*_*d*_ is the probability an individual has the disease given their genetic risk score *G*, calculated as follows:

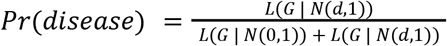

where L(*G* | *D*) is the relative likelihood of genetic risk score *G* with respect to a distribution *D* and *d* is the mean shift in the distribution of genetic risk among cases, ranging from 0.5-1.5 in increments of 0.1. The heritability was calculated using Nagelkerke pseudo-R^2^ in a logistic regression of the phenotype on *G*. We then carried out a logistic regression of simulated phenotype on F_ROH_. We repeated this for 100 simulations, and then calculated power as the fraction of simulations in which the F_ROH_ effect size was positive and its p-value was less than a given cutoff (p<0.05 or p<0.05/61).

## Supplementary note

### Explanation for how autozygosity influences the additive variance of a trait

For illustration, consider a causal locus for a genetically additive trait as is assumed in standard GWAS, where being heterozygous increases risk towards the disease and being homozygous for the alternate allele increases risk twice as much as being heterozygous. Thus, we can code the risk incurred at the locus as 0 for homozygous reference allele, 1 for heterozygous, and 2 for homozygous alternate allele. Assume the locus has risk allele frequency *p*. For an individual not autozygous at the locus, the variance for the coded genotype is equivalent to the variance of a binomial distribution Var 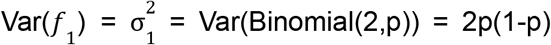. However, for an individual autozygous at the locus, the variance is equivalent to Var 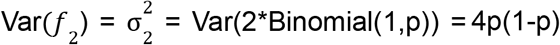.

Given an individual with inbreeding coefficient F (which we approximate by F_ROH_), the variance at the locus of the mixture distribution is:

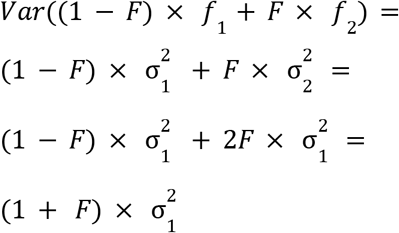

which is equivalent to results of a complementary derivation from Falconer et al.^16^ Thus, extending this argument to multiple risk loci, autozygosity linearly increases variance in risk towards a trait that has an entirely additive architecture. Assuming a liability threshold model, the increased additive variance will lead to individuals with higher F_ROH_ having a greater chance of passing the disease threshold even in the absence of non-additive effects, and may induce an association between F_ROH_ and the trait.

## Supplementary Figures

**Supplementary Figure 1.**
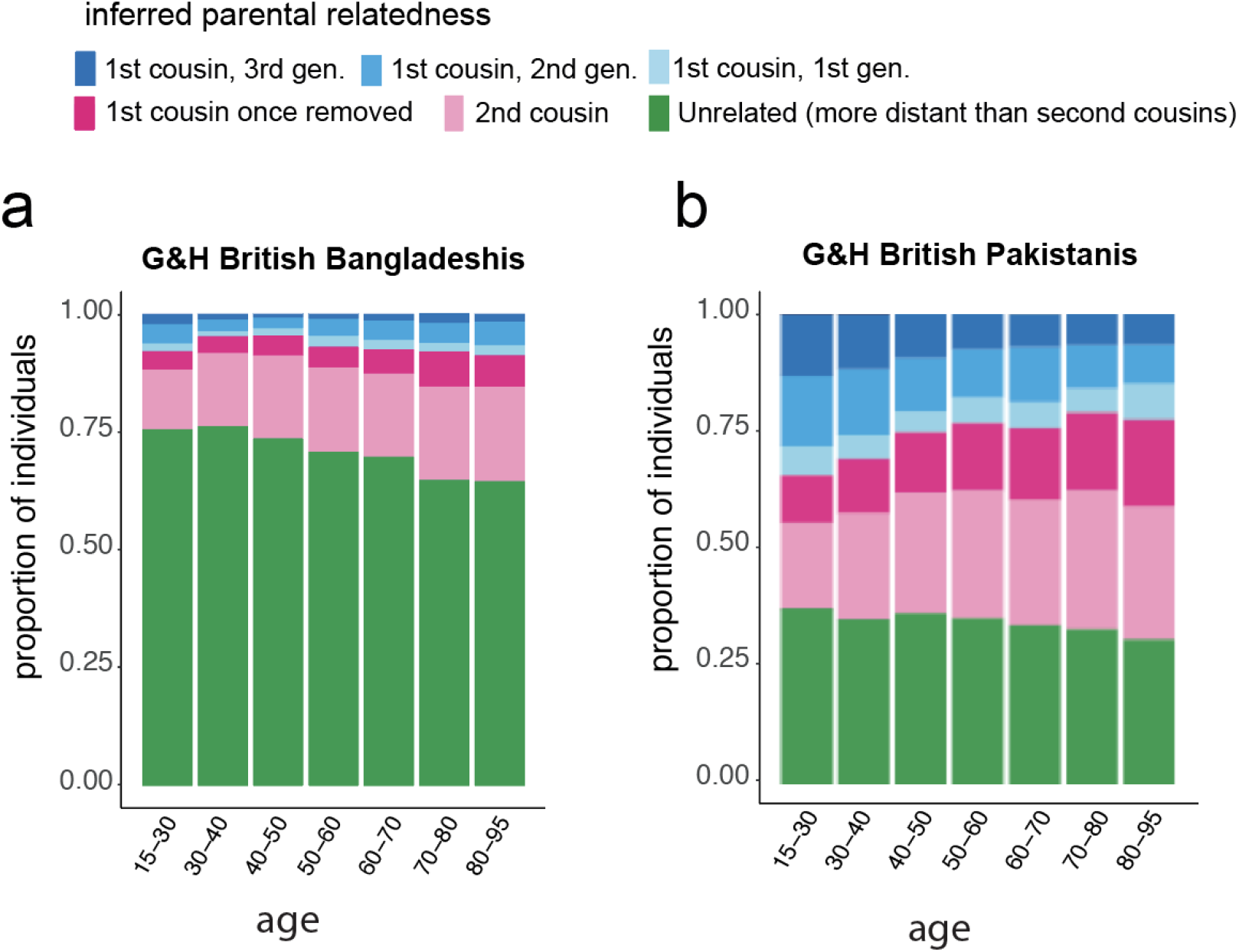
Stacked bar plots showing inferred parental relatedness by age bin in G&H (a) British Bangladeshi individuals and (b) British Pakistani individuals. The inferred categories of parental relatedness include up to three generations of first cousin marriages, first cousin once removed, second cousin or unrelated.

**Supplementary Figure 2.**
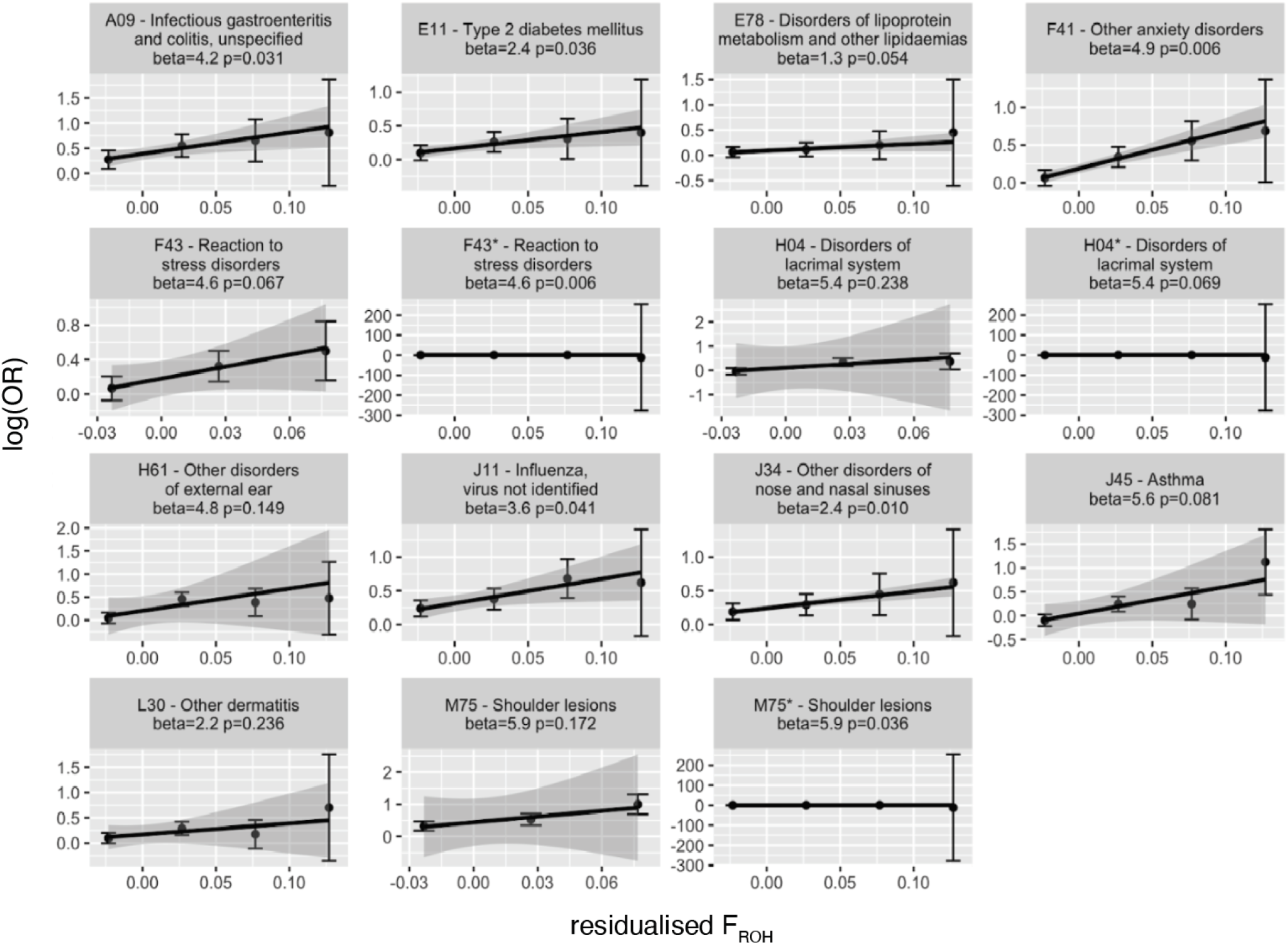
Log(OR) increase in disease risk across residualized F_ROH_ bins by quintiles in the meta-analysis of the highly consanguineous cohorts. The Log(OR) are expressed with respect to the lowest quintile of residualized F_ROH_ values. Error bars reflect standard error (SE) and lines reflect a linear regression of log(OR) on residualized F_ROH_ values with shading representing the SE of the slope. For some traits, the Log(OR) SEs were >200 for the last quintile. For these traits, we plotted them twice, once excluding the last quintile estimate, and once including the estimate, designated with a * following the three letter code. Effect sizes (beta) and p-values are from an inverse-variance weighted linear regression of the log(OR) on the residualized F_ROH_ quintiles.

**Supplementary Figure 3.**
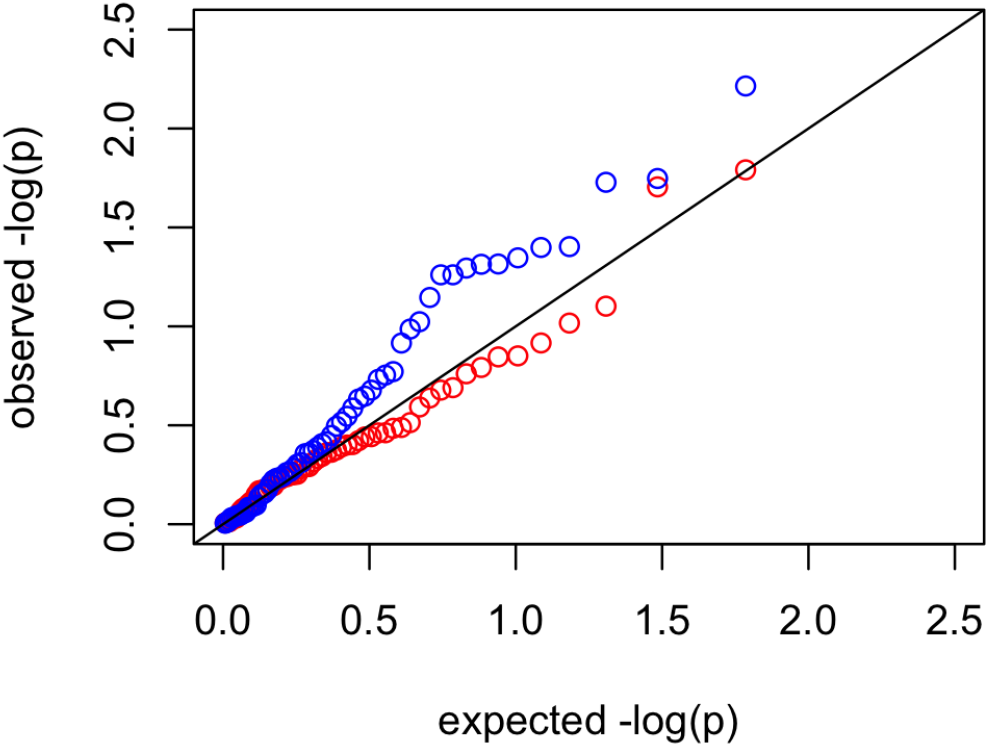
QQ-plot of p-values from a Cochran’s Q test for heterogeneity for all 61 diseases tested across G&H, UKBEUR and UKBSAS, using the full cohorts (blue) and the highly consanguineous cohorts (red). The black line is y=x.

**Supplementary Figure 4.**
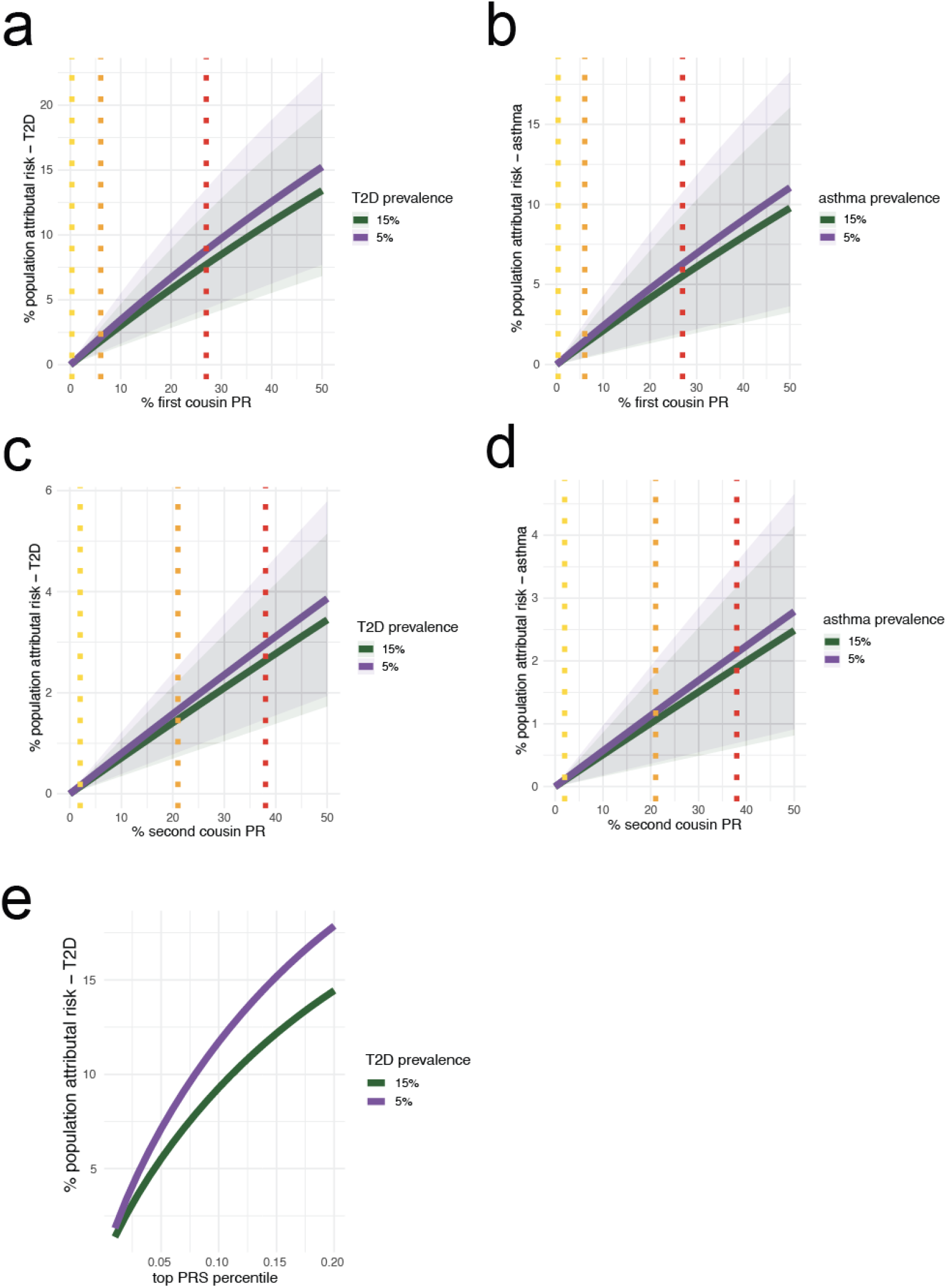
Percent population attributable risk (PAR) for varying degrees of prevalence of parental relatedness for (a), (b) T2D and (c), (d) asthma and (e) for varying fraction of individuals in the top percentiles for T2D polygenic risk score (PRS). (a), (c) show PAR owed to first cousin PR, (b), (d) for second cousin parental relatedness, and (e) for PAR owed to individuals in the top T2D PRS percentiles. Dotted lines indicate the population prevalence estimates for the indicated class of consanguinity in UKBEUR (yellow), G&H British Bangladeshis (orange), and G&H British Pakistanis (red). The prevalence of the disease in the nonconsanguineous populations was used to calculate the RR for each disease using 5% and 15% prevalence, shown in purple and green, respectively. Shaded areas indicate 95% CI for the estimated RR.

**Supplementary Figure 5.**
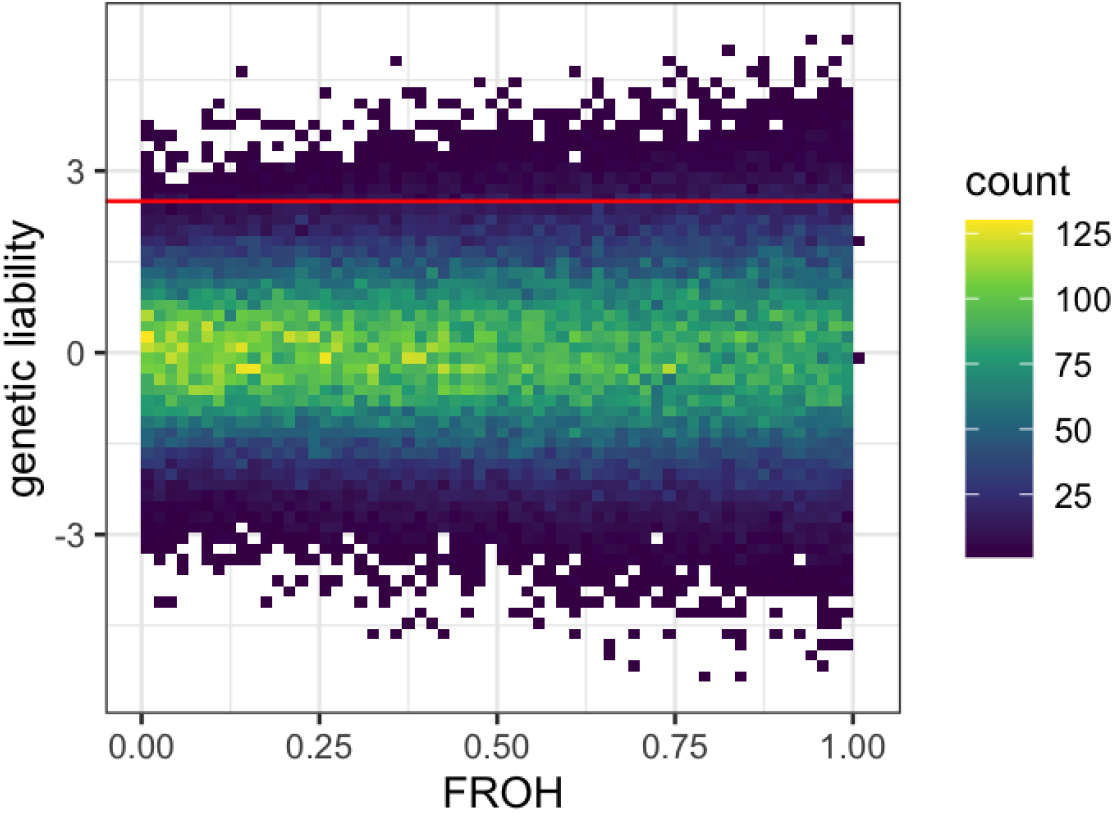
Demonstration of how a correlation between F_ROH_ and disease status can arise in a trait with solely additive genetic genetic architecture. Here, we simulate additive genetic liability and F_ROH_ values for 100,000 individuals. The variance of additive genetic liability towards a trait increases with increasing F_ROH_. If we imagine that individuals with a genetic liability > 2.5 (as shown by the red line) will be disease cases, more individuals will surpass the threshold at higher values of F_ROH_ due to the increased variance in genetic liability. Thus, F_ROH_ could correlate with disease case status when the trait has a purely additive genetic architecture.

**Supplementary Figure 6.**
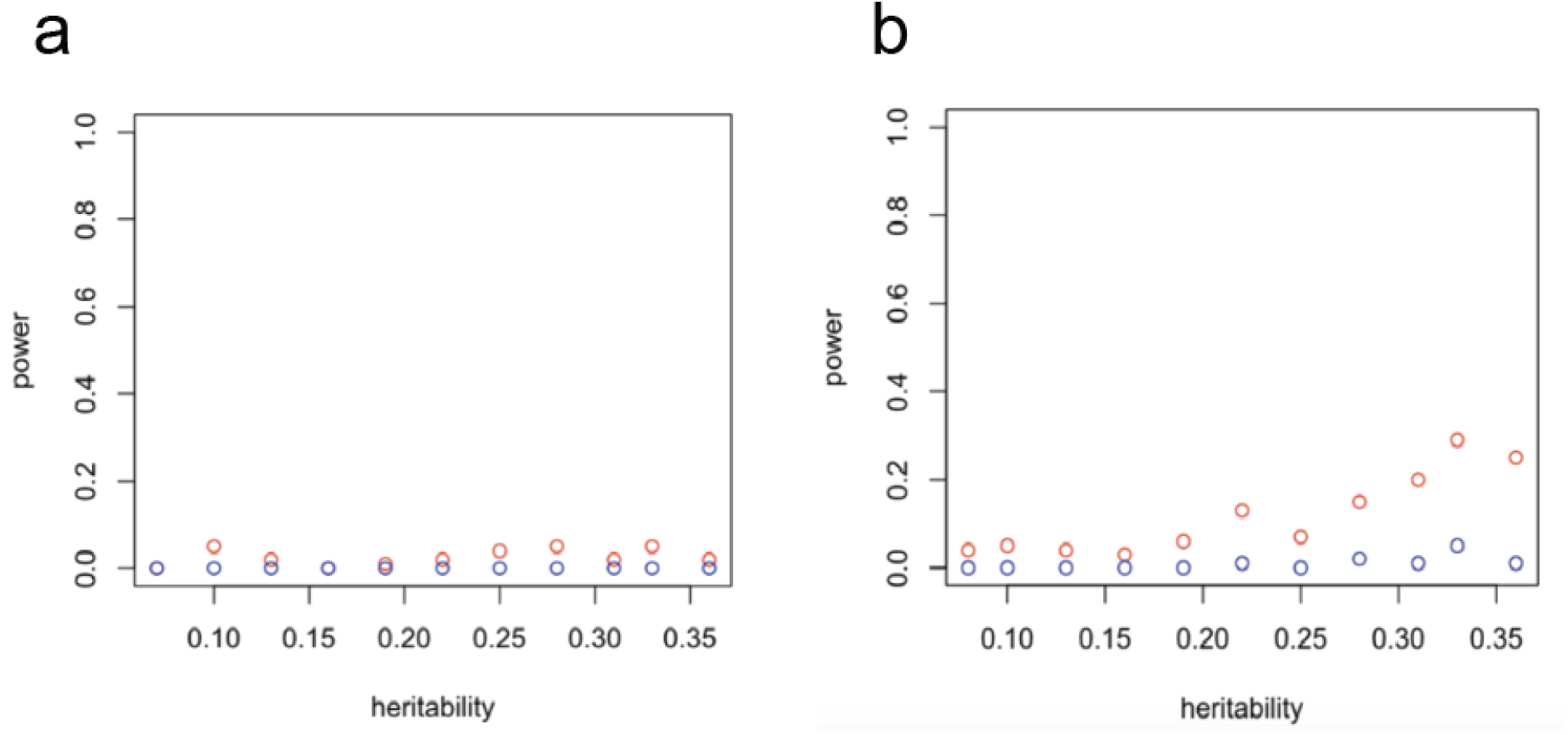
Power to detect significant associations between F_ROH_ and a binary trait with a purely additive genetic architecture and varying heritability. Panel (a) with F_ROH_ values drawn from a lognormal distribution with variance of 0.5 and mean -2.5 and values restricted to be between 0.02-0.18 (i.e. mimicking the observed distribution in Figure 2b) and panel (b) shows the power to detect associations with F_ROH_ values drawn uniformly from 0 to 1. Red is the power for p<0.05 and blue for p<.05/61. Power determined with 100 simulations.

## Supplementary Tables

**Supplementary Table 1**. Associations between F_ROH_ and ICD10 codes in the highly consanguineous cohorts from G&H and UKB. Columns indicate the effect sizes (beta), standard errors (SE), and p-values for the associations in each cohort as well as in the meta-analysis. FDR corrected q-values and Cochrane’s Q test for heterogeneity are shown for the meta-analysis.

**Supplementary Table 2**. Associations between F_ROH_ and ICD10 codes in the full cohorts from G&H and UKB. Columns indicate the effect sizes (beta), standard errors (SE), and p-values for the associations in each cohort as well as in the meta-analysis. FDR corrected q-values and Cochrane’s Q test for heterogeneity are shown for the meta-analysis.

**Supplementary Table 3**. Associations between F_ROH_ and phenotypes in the between-sibling analysis from 23andMe. We show the effect size (beta), standard error(SE) and p-value from a meta-analysis across three chips.

**Supplementary Table 4**. Distribution of F_ROH_ values across cohorts. Per cohort, the quintile distribution of F_ROH_ is shown as well as the mean and standard deviation.

## Supplementary Methods

### Generating ICD-10 codes in G&H by integrating multiple EHR modalities

We derived ICD10 diagnostic codes for each participant with linked healthcare data in Genes & Health. Our methods were designed to closely resemble those used in UK Biobank. For each ICD10 code, we determined whether the participant had any diagnostic codes equivalent to the ICD10 code, the date of the earliest diagnostic code, and the data sources which corroborated the presence of the ICD10 code. In total, we combined data from the following different sources: Barts Health inpatient and outpatient care (native format ICD10, n=23,940 unique pseudoNHS numbers with >=1 code, clinical coding), Barts Health inpatient and outpatient care (native format SNOMED description IDs, n=20,967 unique pseudoNHS numbers with >=1 code, directly coded by healthcare professionals), Bradford Teaching Hospitals inpatient and outpatient care (native format ICD10, n=1,615 unique pseudoNHS numbers with >=1 code, clinical coding), Bradford Teaching Hospitals inpatient and outpatient care (native format SNOMED description IDs, n=1,740 unique pseudoNHS numbers with >=1 code, directly coded by healthcare professionals), primary care observations from the Discovery Clinical Commissioning Group (CCG) and Tower Hamlets (native format SNOMED concept IDs, n=39,077 unique pseudoNHS numbers with >=1 code, coded directly by primary care professionals), NHS Digital Hospital Episode Statistics (both Admitted Patient Care and Outpatient Care), and mortality records (native formats ICD10).

First, we mapped SNOMED description IDs to SNOMED concept IDs for clinician-coded SNOMED codes pertaining to participants who had healthcare encounters at Bradford Teaching Hospitals or Barts Health. The SNOMED mapping file was downloaded from the NHS Digital website on 12/05/22. We used SNOMED build SNOMEDCT2_32.12.0_20220413000001 - the 20th April 2022 minor release (fileset uk_sct2cl_32.12.0_20220413000001Z.zip). This folder contains four separate link files referring to the international SNOMED edition and three distinct UK-specific editions. These files contain mapping for SNOMED descriptionIDs to SNOMED conceptIDs. We collated them into a single mapping reference. All description IDs map onto a single conceptID. This relationship is many-to-one: each descriptionID maps to a single conceptID, but each conceptID can be referred to by several descriptionIDs (the median is three). In total we used a mapping reference consisting of 1,746,657 unique SNOMED description IDs mapping to 578,387 unique SNOMED concept IDs.

For the Barts Health data, we obtained three separate datasets containing records of ‘Diagnoses’, ‘Problems’, and ‘Procedures’ respectively. These files were merged with the mapping files based on the description ID. We excluded codes with a missing SNOMED description ID. Overall we were able to successfully map a high proportion of SNOMED description IDs to concept IDs:

- Diagnoses: 118191 out of 138235 records mapped (85.5%)
- Problems: 31006 out of 31084 records mapped (99.75%)
- Procedures: 3518 out of 3586 records mapped (98.1%)

The most common unmapped code was a code for ‘Venous Thromboembolism Risk Assessment’ (n=13,887 codes), an administrative code of no diagnostic relevance, referring to a standard thromboembolism risk checklist completed on patient admission within Barts Health. Exclusion of this code improved the mapping for the diagnoses dataset from 85.5% to 95.2%. We performed identical mapping for Bradford Teaching Hospitals ‘Diagnoses’ and ‘Problems’ data with a similar successful mapping percentage.

Next, we mapped these codes to ICD10 using the most recent SNOMED maps from NHS digital (SnomedCT_InternationalRF2_PRODUCTION_20210131T120000Z and SnomedCT_UKClinicalRF2_PRODUCTION_20220413T000001Z). We combined the UK and the international map. We restricted this map to SNOMED concept IDs which mapped to a single 3-digit ICD10 code (i.e. a 1-to-1 relationship), resulting in 119,459 individual SNOMED concept IDs. We combined the derived SNOMED concept IDs from step 1 with ‘directly coded’ ICD10 data for each participant in Barts Health and Bradford data separately. 4-digit ICD10 codes were truncated to the first three characters. We then processed data from two primary care networks: the Discovery Clinical Commissioning Group (CCG) network and Tower Hamlets. These data were provided as SNOMED concept IDs and were mapped to ICD10 codes using the same 1:1 mapping approach as for primary care data. Overall between 3% and 8% of all primary care codes were successfully mapped to ICD10 codes, reflecting the large number of administrative and measurement codes recorded in primary care, e.g. ‘text message sent to patient’, ‘blood pressure recording’, and ‘body mass index’.

We then combined 3-digit ICD10 codes derived from these sources (primary care, Barts Health, Bradford Teaching Hospitals) with data exports from NHS Digital (mortality records, HES outpatients and HES APC). Mortality records were searched for underlying cause of death (provided in ICD10 3-digit format). HES-APC codes were used to extract all diagnostic codes recorded during an admission (provided in ICD10 format). We used the admission date as the date of the report. HES-OP data were used to extract all diagnostic codes recorded in relation to the appointment, also provided in ICD10 format. The appointment date was used as the date of report. All ICD10 codes were truncated to 3-digit codes. We excluded ICD10 codes describing generic symptoms rather than disease entities (codes beginning R-Z). For each ICD10 code and each participant, we determined the presence/absence of the ICD10 code (in any health records), the data sources supporting the presence of the code, and the earliest recorded code. When determining the earliest reported code we excluded codes which encode ‘special dates’ in electronic healthcare records (placeholders for missing data) - 1/1/1860, 30/12/1899, 31/12/1899, and 1/1/1900. Similarly to UKB, we derived the ‘source of first report’ field by taking the earliest reported source for the ICD code and specifying whether other data sources supported the code. e.g. if an individual has a diagnostic code for G35 in primary care records and Barts Health data, with the first primary care code being recorded earlier, their ‘source of first report of G35’ value would be ‘Primary care and other sources’. For simplicity, we grouped data sources into ‘secondary care’, ‘primary care’, and ‘mortality’.

Overall, we successfully mapped data for 46,279 unique NHS numbers, 1,926 unique 3-digit ICD10 codes, and 2,976,436 individual diagnoses.

## Notes

### Author Declarations

The London South East NRES Committee of the Health Research Authority gave ethical approval for the G&H work (14/LO/1240, dated 16 September 2014). The North West Multi-centre Research Ethics Committee (MREC) gave ethical approval for UK Biobank. Ethical & Independent Review Services of Salus IRB gave ethical approval for 23andMe research.

