## Supplementary material for "Influence of autozygosity on common disease risk across the phenotypic spectrum": Frequently Asked Questions

### Some questions and answers regarding the paper “Influence of autozygosity on common disease risk across the phenotypic spectrum”

##### 1. Who conducted this study?

The study was conducted by a team of international researchers in the fields of genetics, medicine, and epidemiology. Most of the team are based at the Wellcome Sanger Institute near Cambridge or at Queen Mary, University of London, both in the United Kingdom.

Questions 5 and onwards tell you more about the study and what we found. If you are not familiar with the concepts of DNA, consanguinity or autozygosity, please see Questions 2-4.

#### Background

##### 2. What is DNA? What does it do?

DNA stands for deoxyribonucleic acid. It is a complex molecule that contains information telling organisms (including humans) how to develop and function. DNA is composed of chemicals called “nucleotides” that are linked to each other. There are four different types of nucleotides, referred to by the letters of “A”, “T”, “C”, and “G”. They provide instructions for the cells. Just as we can read sentences, our cells have ways they can “read” the letters in the DNA.

DNA is packaged into large units called “chromosomes”. Each of our cells contains 46 chromosomes that come in 23 nearly-identical pairs. Each parent passes down 23 chromosomes to their child, making up one copy of each chromosome pair. In the cartoon below, you can see an illustration of what your DNA looks like. Each squiggle represents a chromosome and is numbered from 1 to 23. The colours show chromosomes that either come from the biological father in blue or the biological mother in pink, and you can see how they come in pairs. For example, your father gives you one copy of chromosome 1 and your mother gives you another copy.

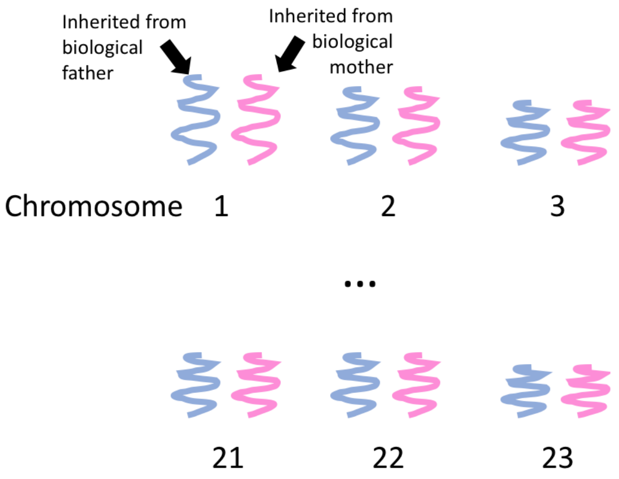


Among humans, the chromosome pairs are mostly identical in terms of the order of the letters in the DNA. However, every person has a slightly different order of letters in their DNA (i.e. genetic variation), which is what makes each person's set of chromosomes unique (see picture below for an illustration). In many cases, genetic variation has no impact on how our cells interpret our DNA, but in some cases it can increase chance towards certain diseases or contribute to traits like height and weight. By understanding how genetic variation impacts disease, we can start to disentangle which aspects of biology are important for the trait.

| | | | | | | | | | |
|---|---|---|---|---|---|---|---|---|---|
| Person 1 | A | T | T | G | C | C | G | G | A |
| Person 2 | A | T | T | G | A | C | T | G | A |
| Person 3 | A | T | T | G | A | C | G | G | A |
| position | 1 | 2 | 3 | 4 | 5 | 6 | 7 | 8 | 9 |

This figure shows an example of genetic variation. Here you see three people's DNA codes in a specific region on one chromosome (e.g. chromosome 1 inherited from the biological father). Here, you can see person 1 has the letter "C" at position 5, while the other two people have a different letter there, "A". This position makes person 1's DNA unique from person 2 and 3. At position 7, you can see that person 2 has the letter "T" while the other two have the letter "G", making person 2's DNA unique. We would say that there is *genetic variation* between people at these positions. There are millions of positions where humans have genetic variation. No two people (except identical twins) have the same order of letters in their entire set of DNA molecules.

#### 3. What is consanguinity? What is autozygosity and why might it be relevant to disease?

Consanguinity is the social and cultural practice of marriage between two blood-related individuals who share a recent common ancestor (e.g. grandparent or great-grandparent). The term is generally used to refer to marriage between people who are second cousins (i.e. sharing a great-grandparent) or closer. Consanguinity is practised all over the world at different rates. In some regions such as parts of North Africa and parts of South Asia, it has been reported that a high fraction (>20%) of all individuals have related parents, while in other regions such as Europe and the Americas, it is less than 2% of individuals. Different cultures have different views on consanguinity, with some seeing it more favourably than others. Rates of consanguinity have also changed across time - for example, it used to be common in Europe until well into the 19th century.

When two people who are closely related have a child, there is a higher chance that some of the DNA the child inherits will be the same in both copies, because it comes from a single common ancestor (see the picture below and description under it for a more detailed explanation). When DNA is the same in both copies inherited from a single common ancestor, this is called "*autozygosity*". For example, for people whose parents are first cousins, typically between around 4% and 15% of their DNA (6.25% on average) is *autozygous* (i.e. the same in both copies). This means that people whose parents are related have a high chance of inheriting the same DNA variant from both parents. Some DNA variants are called "*recessive* variants", meaning they only have an impact on traits (e.g. cystic fibrosis) if they are present in both copies of the DNA.

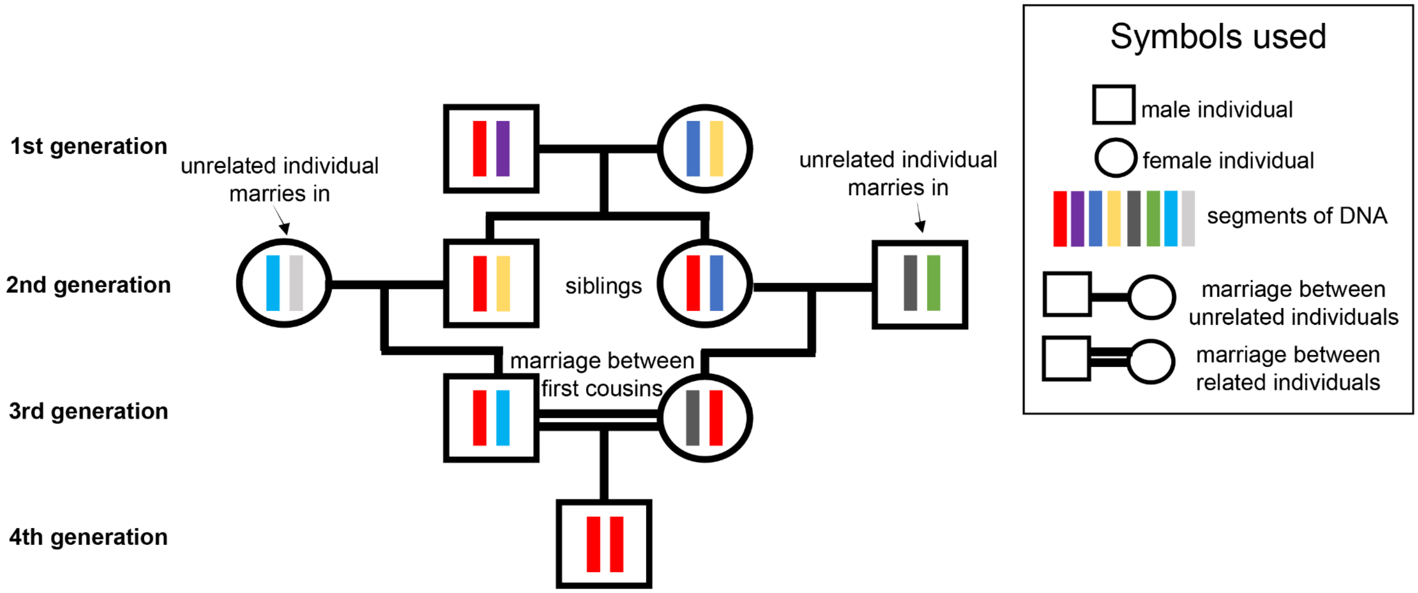


The diagram above shows a family tree for an individual whose parents are first cousins. Biological males are represented by squares and biological females are represented by circles. Horizontal lines connecting the centre of a square to a circle indicate that those two people have a child together, and the vertical lines connect that couple to their child. The colourful bars indicate unique versions of a DNA region that the individuals inherited from their biological mother (circle) or biological father (square). At the top, we see the first generation of people that will be the biological grandparents of the first cousins (third generation) that will have a child together. Each biological grandparent has unique DNA from their biological mother and biological father in this region, shown by the four different colours. The biological grandparents randomly pass down one of the DNA regions to their two children. Here, the biological grandfather passed down the same region to his two children (red version). Each of the siblings then had children with people unrelated to them. The siblings each had children who are first cousins of each other (third generation), and by chance, each of these cousins inherited the same red region of DNA. Lastly, the first cousins had a child (fourth generation). (When people that are related have children, the horizontal lines connecting them are made to be double lines). Again, by chance, each of the first cousins passed down the red version of this DNA region to their child. Now, the consanguineous child is said to be *autozygous* in this region because he received two copies of the red version of this DNA, which originated from a single ancestor, his biological great-grandfather.

##### 4. What was previously known about any health effects of autozygosity?

Autozygosity has been studied in various contexts of human health. For example, it has long been known that people with increased autozygosity have an increased chance of developing rare disorders such as congenital abnormalities (e.g. congenital heart defects, cleft lip) [1,2] and developmental disorders (e.g. severe epilepsy, learning difficulties)[3]. This is because some diseases are caused only by an individual having a particular disease-causing DNA variant inherited from both parents (*recessive* variants). In other words, if an individual only has one copy of this variant then they will not have the disease. It is important to note that even though autozygosity has been shown to increase risk of these rare disorders, the actual number of impacted individuals is relatively small. For example, one study of British-Pakistani individuals found that 2.6% of babies born to unrelated parents had a congenital anomaly compared to 6.2% of babies born to parents who are first cousins [1].

Other studies have looked into relationships (correlations) between autozygosity and a range of other traits such as infertility, cholesterol levels, and decreased lung function[4–6]. Although they found some statistically significant correlations, our study shows that some of their conclusions were less robust, probably because they were driven by correlations between autozygosity and social/environmental risk factors (see Question 12).

For context, many diseases (including type 2 diabetes and asthma) are due to a combination of genetic and social/environmental factors. Some diseases are determined almost entirely by one's genetics, and others much less so. Understanding the genetic influences on diseases, even if they are not entirely due to genetic causes, helps scientists develop preventions and treatments for them. Autozygosity is just one aspect of genetics that may contribute to the chance of developing diseases.

#### About our study

##### 5. Why did you conduct this study?

In this study, we sought to better understand whether autozygosity alters our risk towards more common diseases like diabetes and heart disease. The motivation for this is the need to improve our understanding of the genetic factors causing ill-health. This will help in identifying individuals more likely to develop a medical condition, with the potential to develop interventions (e.g. encouraging lifestyle modification, earlier screening) and new drug therapies in future.

#### 6. Why did you choose to study these groups of people?

We used three groups of people (also known as cohorts) in our study, the Genes & Health (G&H), UK Biobank, and 23andMe cohorts.

Most genetic research has been conducted in people with European ancestries. Some of what has been learned from this has been shown to be transferable to different populations, but there are still major disparities in how well we understand the genetic causes of diseases across different populations. People of different ancestries have different susceptibility to various diseases. For example, hereditary hemochromatosis is more common in European-ancestry populations than other global populations due to differences in genetic backgrounds.[7] In contrast, type 2 diabetes is much more prevalent in South Asian populations compared to people with European ancestries in the UK. This may be due to both genetic and environmental/social factors. A sedentary lifestyle, smoking, and having access to nutritious foods are all known risk factors for type 2 diabetes, which may vary in prevalence between cultures and ancestral groups. However, there's also reason to believe that this difference in type 2 diabetes prevalence may be due at least in part to individuals with South Asian ancestries having a different genetic background. Studies in communities with more diverse ancestries will help us improve risk prediction and improve healthcare for all populations.

The UK Biobank study https://www.ukbiobank.ac.uk/ was established as one of the first large-scale cohorts with genetic and disease data on over 500,000 people. This cohort includes individuals from many ancestry backgrounds, but the majority of individuals are of European ancestries. We focus on individuals with either European or South Asian ancestries from UK Biobank.

The G&H study https://www.genesandhealth.org/ focuses on British Pakistani and British Bangladeshi individuals sampled in London, Bradford and Manchester, and was established to improve our knowledge of health and genetics in these under-studied communities, which make up significant minority populations in the UK. Amongst other things, the data are being used to study the genetic contributions to diseases like type 2 diabetes and heart diseases, which are particularly common in people with South Asian ancestries.

The 23andMe cohort https://www.23andme.com/en-gb/research/ consists of data collected on individuals who purchased 23andMe's personalised genetic testing product and who consented to having their data used in research. It is one of the largest genetic datasets in the world, including over 8 million individuals of diverse ancestries (mostly living in the United States). In our study, we only conducted analyses in individuals who had one or more siblings in the 23andMe cohort.

The UK Biobank+G&H data and the 23andMe data were analysed by separate groups of researchers. 23andMe did not access any data from the UK Biobank or G&H cohorts through our study.

By including ancestrally diverse individuals from these three cohorts, we were able to assess how generalizable our results are and increase our confidence in our findings.

#### 7. What did you find?

In this study, we looked at a wide range of diseases to see whether autozygosity impacts risk of them. Many of these diseases (including type 2 diabetes, asthma and anxiety) are known to be partly due to genetic variants, but it was unknown whether autozygosity impacted them. Briefly, our main findings were:

- Amongst British Pakistani individuals, the rate of individuals with parents who are first cousins is slightly higher in younger than older people. For example, 23% of individuals aged 70-80 were inferred to be offspring of first cousins or closer, compared to 38% of those aged 15-30. However, there was no change in the fraction of individuals with parents who are second cousins or closer.
- Amongst British Bangladeshi and British European individuals, the rate of overall consanguinity (i.e. parents who are second cousins or closer) is (slightly) lower in younger than older people. For example, 37% of British Bangladeshi individuals aged 70-80 were inferred to be offspring of second cousins or closer, versus 25% of those aged 15-30. In British European individuals, 2.10% of individuals between the ages of 60-70 had parents inferred to be second cousins or closer, while for those aged 40-50 it was 1.85%.
- In the two main cohorts we looked at (Genes & Health and UK Biobank), increased autozygosity due to consanguinity is correlated with increased risk for type 2 diabetes, asthma, anxiety-related and stress-related disorders (including post-traumatic stress disorder, PTSD), dermatitis, and certain respiratory and ear disorders. We replicated these findings for type 2 diabetes and PTSD in a separate cohort (23andMe).
- If there were no consanguinity in the British Pakistani population, we estimate there would be 10% fewer people with type 2 diabetes and 8% fewer with asthma (see question 9 for some context). If there were no consanguinity in the British Bangladeshi population, we estimate there would be 3% fewer people with type 2 diabetes and 2% fewer with asthma. These estimates assume that the Genes & Health cohort is representative of the whole British Pakistani/Bangladeshi population with respect to levels of consanguinity.

Although our results are statistically significant, it is important to note that the impact of autozygosity on the chance of developing these diseases is quite low, and that the negative health impacts of consanguinity need to be balanced against positive social impacts. Please see questions 9 and 11 for more detail on this.

#### 8. Tell me more about what you've found and why it's important.

We first looked at how rates of consanguinity differed between groups of individuals with similar genetic ancestries (e.g. British Pakistani-ancestry individuals versus European-ancestry individuals). We used genetic data to infer whether and how people's parents were related. This makes our study's approach different from most of those conducted previously, which typically used self-reported information on consanguinity. As others have seen, we find that British Pakistani and British Bangladeshi people are, in general, more likely to have related parents than European-ancestry individuals. Our study also assessed how these rates have changed over time in the different groups. We found that, amongst British Pakistanis, younger individuals are more likely than older individuals to have parents who are first cousins, but that consanguinity (specifically, having parents who are second cousins or closer) is less common in younger British Bangladeshis than older ones.

We then looked at relationships (correlations) between autozygosity and diseases. We knew there might be factors that we could not account for in the study that make it look like there is a correlation between autozygosity and a disease even if there is no biological causal link between them. For example, hypothetically speaking, if individuals who are more religious were more likely to have related parents than nonreligious people, and religiosity decreases the chance that people will be diagnosed with depression, we might see a negative correlation between autozygosity and depression, even though there is no biological link leading to that correlation. We developed a method to address this issue and showed that it works well (see Question 12 for an explanation of the method). We also found that previous studies that looked for correlations between people's degree of autozygosity and different traits probably did not properly account for this issue.

We then applied our method to look at correlations between people's degree of autozygosity and diseases. We found that increased autozygosity is correlated with a higher chance of having type 2 diabetes, asthma, and anxiety-related and stress-related disorders (including PTSD), as well as other common diseases using the G&H and UK Biobank cohorts (see bullet points above under Question 7). We then found that the results for type 2 diabetes and PTSD replicate in the 23andMe cohort using a different analysis technique (Question 12 explains this technique).

We estimated how much consanguinity changes the rates of having type 2 diabetes and asthma in different populations. We estimated that if there were no consanguinity, the number of people with type 2 diabetes amongst British Pakistanis would be reduced by 10%. This number was lower in British Bangladeshi individuals (3%) and even lower in British-European ancestry individuals (<1%), because they have lower rates of consanguinity. To estimate this quantity, we need to know the strength of the relationship between autozygosity and the disease (estimated in our study), the prevalence of consanguinity in the population of interest (estimated in our study), and the prevalence of the disease in individuals with unrelated parents in the population (unknown, so we varied it between 5% and 15% which spans the probable range of the prevalence in the population from known studies). An assumption of this analysis is that the prevalence of consanguinity within our sample is representative of the wider population of the relevant group (e.g. G&H is representative of British Pakistanis).

Overall, our study contributes to research efforts to understand the genetic causes of disease and to motivate future studies to look at specific types of genetic variation that may impact the diseases we found are correlated with autozygosity. These future studies may help to identify better therapies for these diseases.

#### 9. So are you saying that consanguinity is the reason type 2 diabetes is more common in certain populations?

No, we are not saying that. We found that consanguinity was a relatively small factor, explaining about 10% of the prevalence of type 2 diabetes in British Pakistanis and about 3% in British Bangladeshis. This is a lot less than is explained by other risk factors; for example, a study in the Netherlands found that 71% of type 2 diabetes was explained by modifiable risk factors such as smoking and body mass index[8]. Thus, although consanguinity does contribute to the higher rates of type 2 diabetes in British Pakistanis in particular, there are probably other factors including environmental and socio-economic factors, and more research is needed on these.

#### 10. I have/am at risk of type 2 diabetes. What do these findings mean for me?

The risk of a person developing Type 2 diabetes (T2D) is linked to both genetic and environmental/social risk factors. This paper shows that autozygosity contributes to the genetic risk of T2D. Although we cannot change our genetic risk because it is fixed permanently when we are conceived, a person at risk of T2D can reduce their chance of developing the condition by changing some of their other risk factors, e.g. how active they are or what foods they eat. If you would like to learn how to reduce your risk of T2D, we would recommend consulting the Diabetes UK website. They also have advice on putting T2D into remission.

#### 11. Are you saying that consanguinity is a bad thing because it increases the risk of some diseases?

No, we are not saying that. Although we show strong evidence suggesting that consanguinity does increase the risk of some common diseases (e.g. type 2 diabetes, asthma), **the health risks need to be balanced against social benefits of consanguinity such as stronger family support networks and stronger cultural and community bonds.** The aim of our study is not to pass judgement on the social practice of consanguinity, but rather to guide scientists as to what kind of studies to conduct in the future to better understand these diseases and the biology underlying them, to help develop treatments.

#### 12. What is the approach you developed?

We developed an approach to make sure that correlations between autozygosity and traits or diseases are not simply due to cultural/environmental factors that are correlated with consanguinity, but rather due to genetic effects. The approach involves focusing the analysis on individuals who are offspring of first cousins (whom we refer to as “highly consanguineous” in the paper). Within this group of people, the amount of each person’s DNA that is autozygous (the same in both copies) is typically between about 4% and 15%, but the precise amount is randomly determined, and is not correlated with environmental/social factors. Thus, focusing on this group avoids finding spurious correlations between autozygosity and traits that are actually due to cultural/environmental factors correlated with consanguinity which would be present if we included non-consanguineous individuals in the analysis.

To test how well this approach worked, we looked at correlations between autozygosity and several cultural and environmental factors, namely religiosity, the level of deprivation, educational attainment, and tobacco and alcohol use. When we looked at everyone in the study, we found that autozygosity was significantly correlated with all these factors. However, when we restricted to the set of people we inferred to be the offspring of first cousins, these correlations completely disappeared. (For example, tobacco use was negatively correlated with autozygosity when using all individuals, but not correlated with it at all amongst the offspring of first cousins.) This suggests that the correlation observed when including all individuals was not driven by genetics, but rather was driven by social/cultural/environmental factors that are correlated with having parents who are first cousins. Thus, we think our method of restricting to the children of first cousins works well to remove unwanted correlations between autozygosity and culture/environment. This makes us much more confident that the correlations we saw between autozygosity and diseases within the group of people inferred to be offspring of first cousins are due to true genetic effects.

#### 13. What is the technique you used to analyse data in the 23andMe cohort?

Between siblings that have the same biological parents, the amount of autozygosity naturally varies somewhat by random chance. Siblings share very similar home environments and have the same ancestral background. We looked for relationships between the difference in autozygosity between siblings and differences in whether or not they have a given disease. For example, we looked to see if the sibling with the higher degree of autozygosity in their family is more likely to have the disease than their other siblings, and averaged these results across thousands of families. This technique is typically thought of as being very strong evidence that the difference in genetics is causally related to disease.

#### 14. What are the limitations of this study?

As with nearly every study that uses human cohorts, we are not able to have controlled experiments where we can definitely say that one variable causes a change in another. What we find are correlations between the two variables (in this case, autozygosity and various diseases). In this study we tried our best to make our analyses mimic an experiment, by making use of the random variation in autozygosity amongst individuals who are offspring of first cousins (see Question 12 above). However, we cannot completely rule out that within this group of people, there could be social/environmental factors correlated with autozygosity which are causing the correlations with diseases. However, our analysis of suspected confounders (religiosity, the level of deprivation, educational attainment, and tobacco and alcohol use) suggests that these are not driving the correlations with these particular confounders. There may be other confounders we have not measured - we only analysed the suspected confounders for which we had data in UK Biobank.

Similarly, our study is based only on the cohorts we had access to, and may not generalise to the rest of the population that wasn't represented in these cohorts. For example, the observed changing rates of consanguinity can only be interpreted within the context of the British South Asian individuals included in the Genes & Health cohort (i.e. British Pakistani and Bangladeshi individuals primarily from east London) rather than all South Asians in the UK or the populations in Pakistan and Bangladesh.

A limitation of our results is that we have not identified the specific DNA variants that are driving the associations between autozygosity and diseases. This will require much larger studies than are currently available, and would help us understand the biology of diseases better, and potentially discover new therapies.

Another limitation of our study is that we have only studied individuals with ancestries reflecting broadly two continental groups (European and South Asian) and only from within the UK. Future studies in all of genetics need to continue expanding their cohorts to include individuals with different ancestries. This will help both in making sure our studies reflect the genetic variation in the world and ensuring that our results are generalisable to more individuals.
